# Modeling COVID-19 outbreaks in United States with distinct testing, lockdown speed and fatigue rates

**DOI:** 10.1101/2021.01.04.21249231

**Authors:** J. C. Macdonald, C. Browne, H. Gulbudak

## Abstract

Each state in the United States exhibited a unique response to the COVID-19 outbreak, along with variable levels of testing, leading to different actual case burdens in the country. In this study, via per-capita testing dependent ascertainment rates, along with case and death data, we fit a minimal epidemic model for each state. We estimate infection-level responsive lockdown entry and exit rates (representing government and behavioral reaction), along with the true number of cases as of May 31, 2020. Ultimately we provide error corrected estimates for commonly used metrics such as infection fatality ratio and overall case ascertainment for all 55 states and territories considered, along with the United States in aggregate, in order to correlate outbreak severity with first wave intervention attributes and suggest potential management strategies for future outbreaks. We observe a theoretically predicted inverse proportionality relation between outbreak size and lockdown rate, with scale dependent on the underlying reproduction number and simulations suggesting a critical population quarantine “half-life” of 30 days independent of other model parameters.

## 1. Introduction

The COVID-19 pandemic began in Wuhan, China and spread rapidly throughout the world in early 2020 provoking a wide range of interventions. China gained rapid control of its epidemic, likely due to it’s rapid and strict lockdown as discussed in several studies [4], [7], [15], in contrast to several other countries with slower and less unified efforts. In particular, the United States employed a heterogeneous response, with different scales of quarantine measures (stay at home orders and business closures), despite the first reported case of COVID-19 occurring on January 21, 2020 in Washington state and cases being reported in all 50 states by mid-March [13]. Furthermore, each state displayed distinct exit strategies and lockdown fatigue even though cases never were brought down to low levels. Another feature of the US response was variable testing through time and by state, growing from sparsely to more widely available tests, leading to distinct increasing trends of case detection.

Each state represents distinct realizations of how different response characteristics and case ascertainment correlated to true case burden in the United States, which provides fertile ground for testing outbreak containment strategies. However, successfully fitting an epidemic model to this heterogeneous response exhibited in the first wave COVID-19 outbreak in the US presents a number of challenges. The multitude of epidemic curve shapes induced by the distinct dynamic behavioral and government interventions can lead to issues of over-fitting or a large number of parameters that blur the most important factors [8]. In addition, the unknown true number of cases calls for methods to incorporate testing, mortality or other sources of data (e.g. seroprevalence studies), which is complicated by changing quantities, such as detection [1], [12], [11], or antibody levels [12]. Here we focus on a unified model both simple and flexible enough to fit the wide range of COVID-19 outcomes in the US (through May 31, 2020), which incorporates the critical factors in epidemic trajectory. Furthermore, we construct an appropriate per-capita testing dependent ascertainment rate calibrated with mortality data to both allow more accurate model fits and provide a means of estimating the actual number of cases.

The wide range of quarantine responses in the different states and resultant outcomes allow us to analyze potential responses to future outbreaks utilizing correlation and sensitivity analyses as well as counterfactual simulation studies. In this way, our minimal number of identifiable parameter values can be compared among the states representing crucial control quantities, such as lockdown speed and fatigue, and via analytically derived relations, we link outbreak size as inversely proportional to population quarantine rate. We also provide estimates of commonly used quantifiers of outbreak severity, epidemic trajectory and effectiveness of control measures, such as Infection Fatality Ratio (IFR) and the ratio of (estimated) true cases to reported cases, accounting for common sources of error in their prediction [14], [6]. By estimating these quantities for all states and territories under one modeling framework we provide a comprehensive overview of the first wave outbreak in the United States. Ultimately we synthesize these results into a range of potential future responses which highlight the critical nature of widespread, swift quarantine measures, with a model suggested critical duration, for a rapidly spreading outbreak. These results can be used to inform management strategies both as COVID-19 vaccine is not yet widely available, and for future outbreaks of emergent pathogens.

## 2. Model

First we formulate an epidemic model which can account for the heterogeneous response to COVID-19 outbreaks exhibited in the United States. In [12], we developed an SIR-type model applied to the outbreak in China incorporating terms for responsive self-quarantine (lockdowns) and contact tracing, where the rate of both control actions depend upon current infection rates. Here we focus our modeling framework to fit the wide range of reactionary lockdown measures and testing observed in different states during the first wave of COVID-19. Consider the following system (see also Fig. 1) for susceptible (*S*) and quarantined (or socially distanced) susceptible (*S*_*q*_) population, and infected (*I*) individuals whom ultimately progress to reported (*R*), unreported (*U*), or dead (*D*) cases.

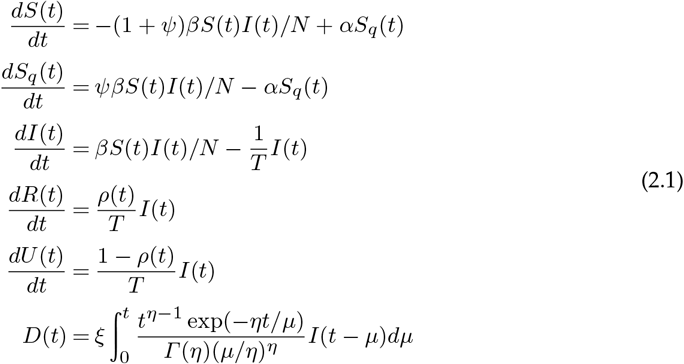

**Figure 1:**
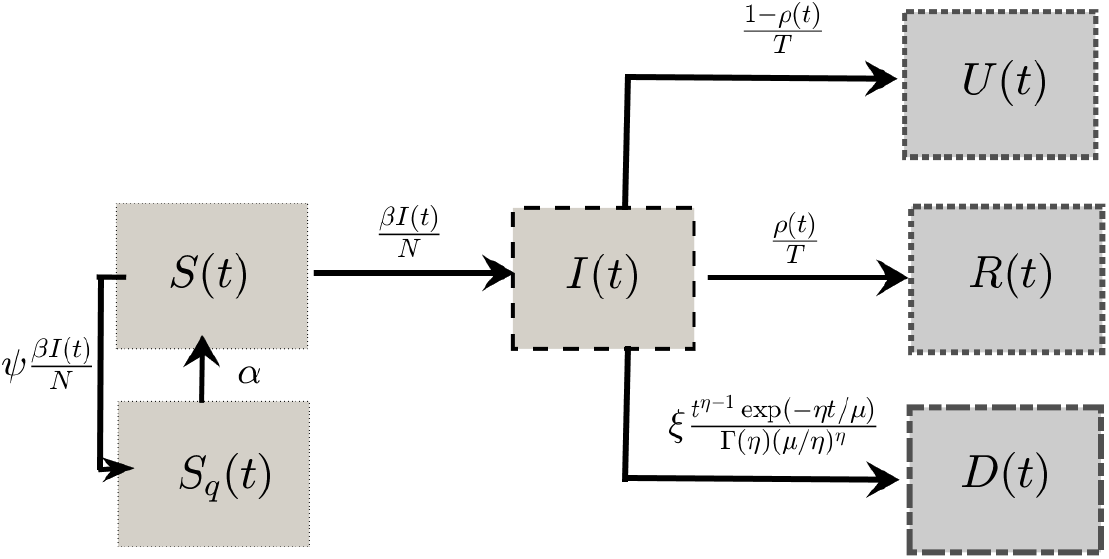
Model compartments are Susceptible Individuals (*S*), Self-Quarantined Individuals (*S*_*q*_), Infected Individuals (*I*), Reported Cases (*R*),Unreported Cases (*U*), and Deaths (*D*). See also table 1

Our model assumes susceptible individuals become infected relative to force of infection 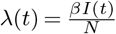, where *β* is the transmission rate and *N* is the population size. In addition susceptible quarantine acts proportionally to force of infection *λ*(*t*) with lockdown (rate) factor *ψ*, as a proxy for the population (or government mandated) reaction to incidence by self-quarantine. While individuals are in quarantine it is assumed that they do not contact with infected population, and that individuals exit quarantine with rate *α*, which we also call *lockdown fatigue*. Infected individuals either have their cases reported (model compartment R), or fail to have their cases reported (model compartment U), with mean infectious period *T* = 7.5 days, with the proportion of individuals in each compartment determined by ascertainment rate *ρ*(*t*), or the proportion of true cases at a given time captured by testing. Regardless of report status death is expected to occur with Infection Fatality Ratio (IFR), represented by model parameter *ξ* after a mean delay of *µ* = 21 days after infection. The model parameters and descriptions are also listed in table 1. In addition to the fixed parameter values for mean infectious period *T* and mean time until death *µ*, we fit the remaining parameters of the model as described in next section, obtaining good fits for the data from all 50 states, Washington D.C. Guam, Puerto Rico, the Northern Mariana Islands, and the US Virgin Islands (see Figure 7 and the SI figs. S1-S14).

**Table 1:**
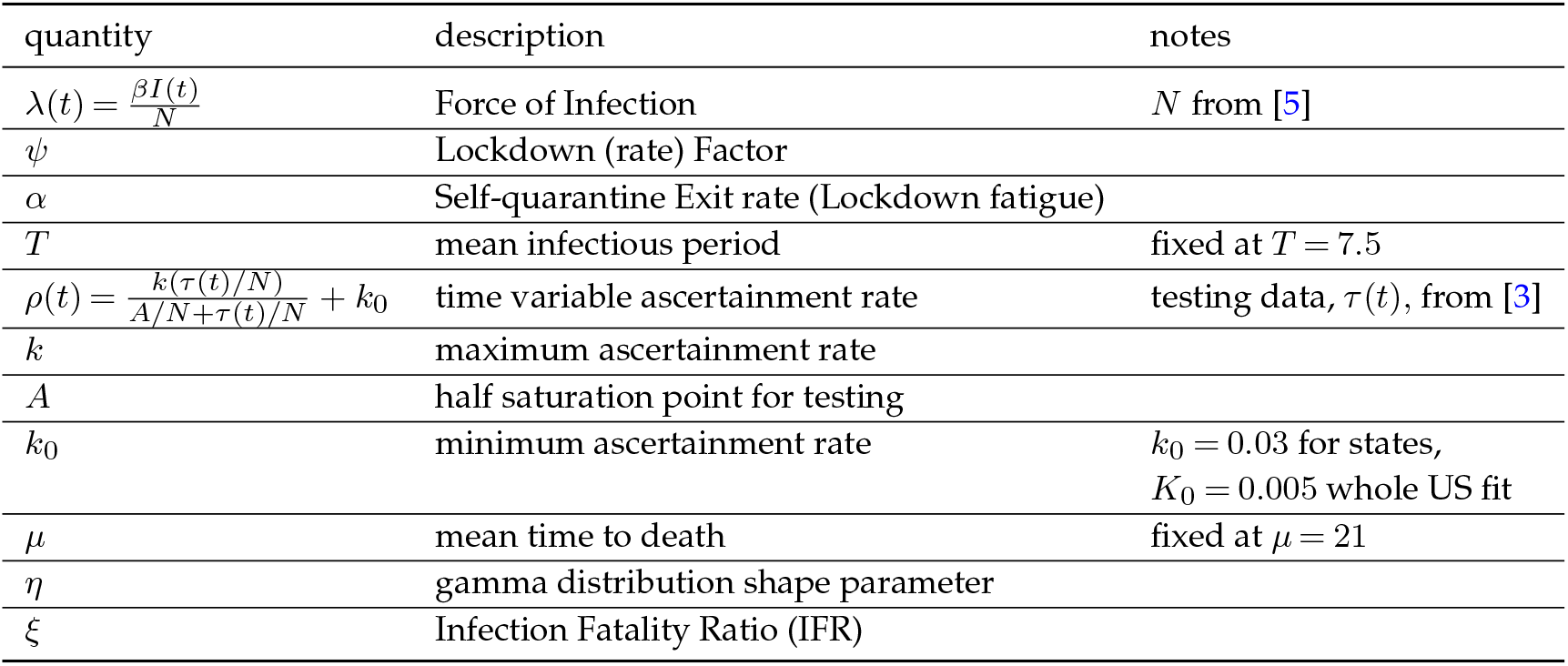
Key Model parameters and quantities with description

## 3. Methods

We now describe the methods of our fitting procedure and analysis of the model. All relevant data and code are available at [9]. Where shown, indicated stay at home orders are based upon [2]. Population levels are from US Census Bureau 2019 projections [5]. To obtain parameter estimates for our model in each state and territory, we utilized cumulative death and case totals [10] as well as daily testing data, inferred from [3]. The data used for the whole US fit was obtained by combining data from all subsidiary states and territories due to concerns about data reliability at the federal level [16]. Even at the state level the raw testing data contained a number of reporting irregularities, as evidenced by several days with negative daily testing instance. To account for this we smoothed the data by averaging with nearest neighbors in such a way as to not change the cumulative testing totals prior to finding the moving average.

We define the ascertainment rate function *ρ* (t) as

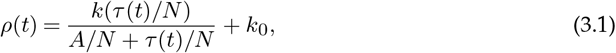

where *τ* (*t*) is the three day moving average of the raw daily testing totals, with parameters *k* as maximum ascertainment rate, *A* as half saturation point in terms of tests, and *k*_0_ as minimum ascertainment rate. Observe that *ρ*(*t*) is chosen to be a saturating function of daily tests as a percent of population (see Figs 2 (b)-(d) and 4 (e). As the number of tests increased, so did the ascertainment rate because the increase in tests was not driven by more demand solely based on rising case numbers, but wider availability and convenience leading more individuals overall to take the tests. The negative concavity of *ρ* as a function number of tests reflects the principle of diminishing returns associated with wait times, and also a larger influence of demand due to actual rising cases once tests become widely available, which together lead to saturation of *ρ* to a maximum level. It is assumed even absent or at low testing, there is a minimal ascertainment rate calibrated based upon low ascertainment levels during the early stages of an outbreak with bounds of .005 and .03 for the whole of the United States and its territories, and for each state or territory respectively. This difference in lower bound was chosen because, as indicated by Figure 5a, many states’ outbreak began significantly before their first reported case, thus when considering the United States as a whole, few cases were ascertained in the initial phase of the outbreak. For each state and territory fit and the whole United States fit, the maximum ascertainment rate was fixed based upon the maximum daily test count as a percent of the states population. This bound was set based in part on the estimated cumulative case total for Connecticut and New York City provided in [12]. The half saturation constants were fit to each state, calibrated by the simultaneous fit of mortality and reported cases, to reflect state specific relationships between testing and ascertainment (see Fig. 2). The obtained ratios of total to reported cases (see table 2) are in line with CDC estimates [1].

**Figure 2:**
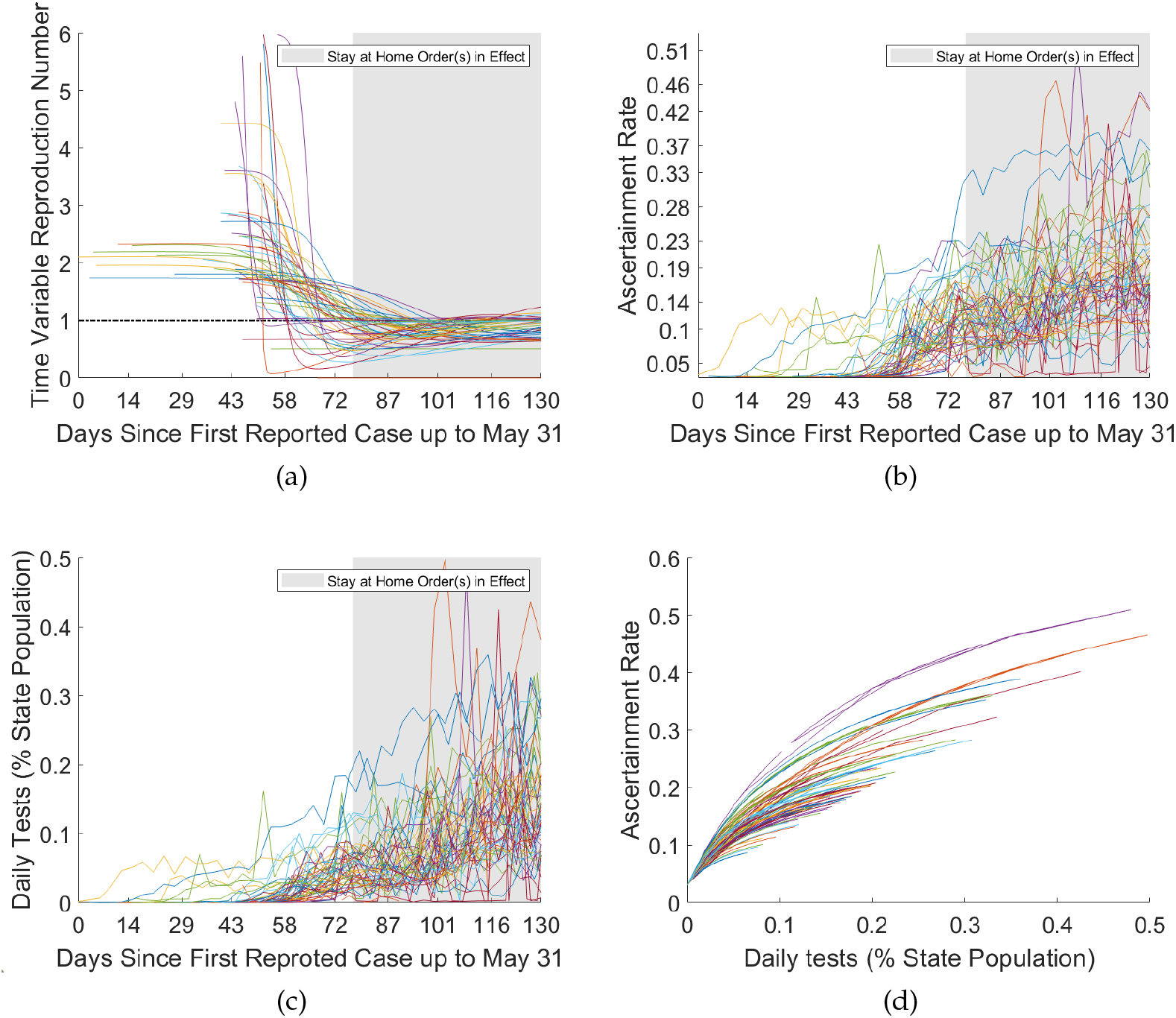
(a) Time variable Reproduction Number, ℛ_*e*_(*t*). States with later reported first case have higher basic reproduction number ℛ_0_ = ℛ_*e*_(0) (b) Time variable ascertainment rate (c) Tests as percent of individual states population (d) Ascertainment rate as variable of daily tests. Ascertainment saturates as daily tests increase.

We numerically solved the non-linear least squares problem, using the interior-reflexive Newton method as implemented by MatLab’s lsqcurvefit function, minimizing objective function

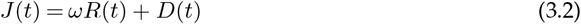

where the ratio of cumulative deaths and reported cases at end-times, 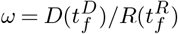, was the chosen positive weight for each state. For each state or territory the time interval considered was from the first confirmed case up until 31 May 2020 for cases, by which time most state’s mandated stay at home orders had ended [1], and June 21, 2020 for deaths. For our fittings the mean infectious period was fixed at, *T* = 7.5 and the mean to death at *µ* = 21 to improve model identifiability. These values are in line with the weighted (by proportion of total cases) mean parameter values obtained from fitting the model without fixing these parameters (see program files at [4]). All other model parameters, as well as *I*_0_, the initial case total, were fit for each state or territory. The fit values (see table 2 and the SI for all parameter values) were then used to provide approximate cumulative unreported case totals (model compartment *U*) and true cumulative case totals (sum of model compartments *U* + *R*). We additionally checked if further model parameters could be fixed while retaining model robustness (see section 1 of the SI), but found that fitting both case ascertainment, *ρ*, and infection fatality ratio, *ξ* (bounded within a feasible range from .001 to .05), were necessary to obtain good fits across all states and territories.

In order to compare % *model predicted positives* (MPP) with % *positives inferred from the data* (PID) these quantities were plotted together with *τ* (*t*) for each state and territory,

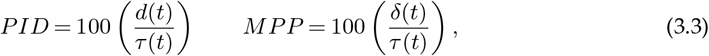

where *d*(*t*) is the three day rolling average of case totals inferred from reported cumulative case totals after applying the same smoothing to daily case totals as was applied to daily testing incidence, and *δ*(*t*) is the daily reported case incidence inferred from the fit cumulative case totals (see Figures 4, 7).

For all states and territories, with the exception of the Northern Marianas Islands (due to that this territory’s fit parameters indicating that all cases were imported), the relationship between model parameters and outbreak trajectory were assessed. The reproduction number ℛ_0_ can be utilized as a single parameter. Indeed, for model (2.1),

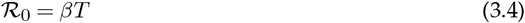

(noting that the susceptible quarantine does not affect ℛ_0_ because it is proportional to force of infection). Then since *T* is fixed (at 7.5 *days*), the quantity ℛ_0_ can be identified with parameter *β*. We computed time and test variable ascertainment rate, *ρ*(*τ* (*t*)) as well as time variable reproduction number, ℛ_*e*_ = (*S/N*) ℛ_0_ (see Figure 2). We also analyzed the association of estimated parameter values with outbreak measures, along with correlation between parameters, using Spearman’s rank correlation (see Figure 5 and SI figure S19 for statistically significant results).

In order to assess identifiability and confidence in our fitting procedure, we conducted uncertainty analysis. For the whole United States and its territories, the uncertainty quantification was carried out in the following manner:

i. Simultaneously fit the model (2.1) to cumulative case and death totals, and national testing data as described above.
ii. Obtain the inferred fit daily case total curve, and under the assumption that the reporting error is normally distributed and relative in magnitude to the reported total at each data point:

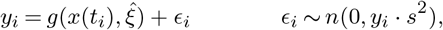

where *g* is the true number of daily cases and 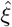 the set of true parameter values. We generated 10,000 cumulative case data sets with noise level of 60%, we then summed these daily case results and refit the model to each of them and the original death data simultaneously. For the results in table 2,3 and Figure 4 a value of *s* = .6 was used because this value causes the synthetic data-sets to cover the original data (see the SI figure 18).
iii. Arrange the generated values in increasing order and remove the top and bottom 2.5% in order to obtain the desired approximate 95% confidence intervals.

The resulting fit, confidence intervals, and average relative errors (see Figure 4 and table 3) indicate that the key model parameters are all practically identifiable assuming the 60% noise level in daily case reporting.

In addition, to further examine the relationship between *ψ, ℛ*_0_, *α* and outbreak trajectories, we also highlight several states with different relative relationships between *ψ* and *α* representing the range of outbreak responses in the United States, and conduct both sensitivity analysis and counterfactual simulation studies with these examples (see Fig. 7). We fixed model parameters as those obtained from our fitting process, and varied ℛ_0_,*ψ* and *α* to examine their differential impacts on epidemic trajectory. The fit plots for all states and territories are included in the SI.

## 4. Results

Our unified model simultaneously fits each state’s cumulative mortality and reported case data, along with daily percent positive tests through our calibrated ascertainment rate, starting at first reported case to May 31, 2020 (as described in Methods). The estimated parameters, particularly quarantine (rate) factor, *ψ*, lockdown exit rate (fatigue), *α*, reproduction number, ℛ_0_, and Infection fatality ratio, *ξ*, provide key information about the variability of COVID-19 spread and response in the United States. The resulting parameter values (table 2) and associated fits provide true (reported and unreported) cumulative case estimates for all 55 states and territories considered. These range in value from as low as 0.42% of population (Hawaii) to as high as 10.41% of population (New York State) over the time period, with an estimated 3.51% of the entire US population having been infected as of May 31, 2020 (*Confidence Interval* 2.72-4.43% population). These values represent both the wide range of observed outcomes and highlight the importance of incorporating testing data to obtain sufficient model flexibility (Fig. 2, see also SI figures S15,S16 and S4-S16). Indeed, the ratio of true cumulative cases to reported cases across states ranged from as low as low as 2.99 (Rhode Island) to as high as 17.88 (the US Virgin Islands) with a value of 6.62 (*CI* 5.54-8.20), in line with CDC estimates [3]. Our fit IFR, both for every state and for the entire United states, works to counter both a common source of overestimation and a common source of underestimation [14], [6]. Indeed, our gamma distributed delay (mean *µ* = 21) of days after infection until death and fitted true cumulative case totals with ascertainment rate *ρ*(*t*), together estimate IFR ranging from as low as 0.001 (Guam) to as high as .0156 (Rhode Island). There were outbreaks from mild to severe toll; with the fit IFR for the United states being 0.009 (*CI* .007 - .011) as of June 21 (cut-off date for deaths associated with infections on or before May 31), similar to estimates from a different retrospective study [14].

To further dissect the relationships between model parameters and estimated quantities we next turn to correlation analysis. The following notable pairs of epidemiologically and statistically significant variables were found: *I*_0_ vs date of first reported case (after Jan 21) with Spearman correlation = .48 (*p* = 2.3 × 10^−4^); mortality (% population) vs. *ψ* with = ϱ −.53 (*p* = 3.7 × 10^−5^; true Case Estimate vs. *ψ* with = ϱ −.68 (*p* = 2.1 × 10^−8^); *ψ* vs. *α* with ϱ = .52 (*p* = 5.8 × 10^−5^; peak daily cases vs. *ψ* with = ϱ −.72 (*p* = 1 × 10^−9^; time to peak daily cases vs. ℛ_0_ with = ϱ −.34 (*p* = 1.6 × 10^−2^; cumulative case estimate vs. IFR, = *ϱ* .33 (*p* = 1.6 × 10^−2^); cumulative reported cases vs. IFR with = .71 (*p* = 4.7 × 10^−5^); cumulative case estimate/reported cases vs. IFR with *ϱ* = −.63 (*p* = 2.8 × 10^−7^. This analysis (see Figure 5 and SI figure S19 for all correlation plots) together with our three variable scatter plots in Figure 3 suggest that *ψ* dominates *α* and *R*_0_ in terms of statistically significant correlation between model parameters and quantities of interest such as cumulative and peak daily case total as well as mortality and lockdown fatigue.

**Figure 3:**
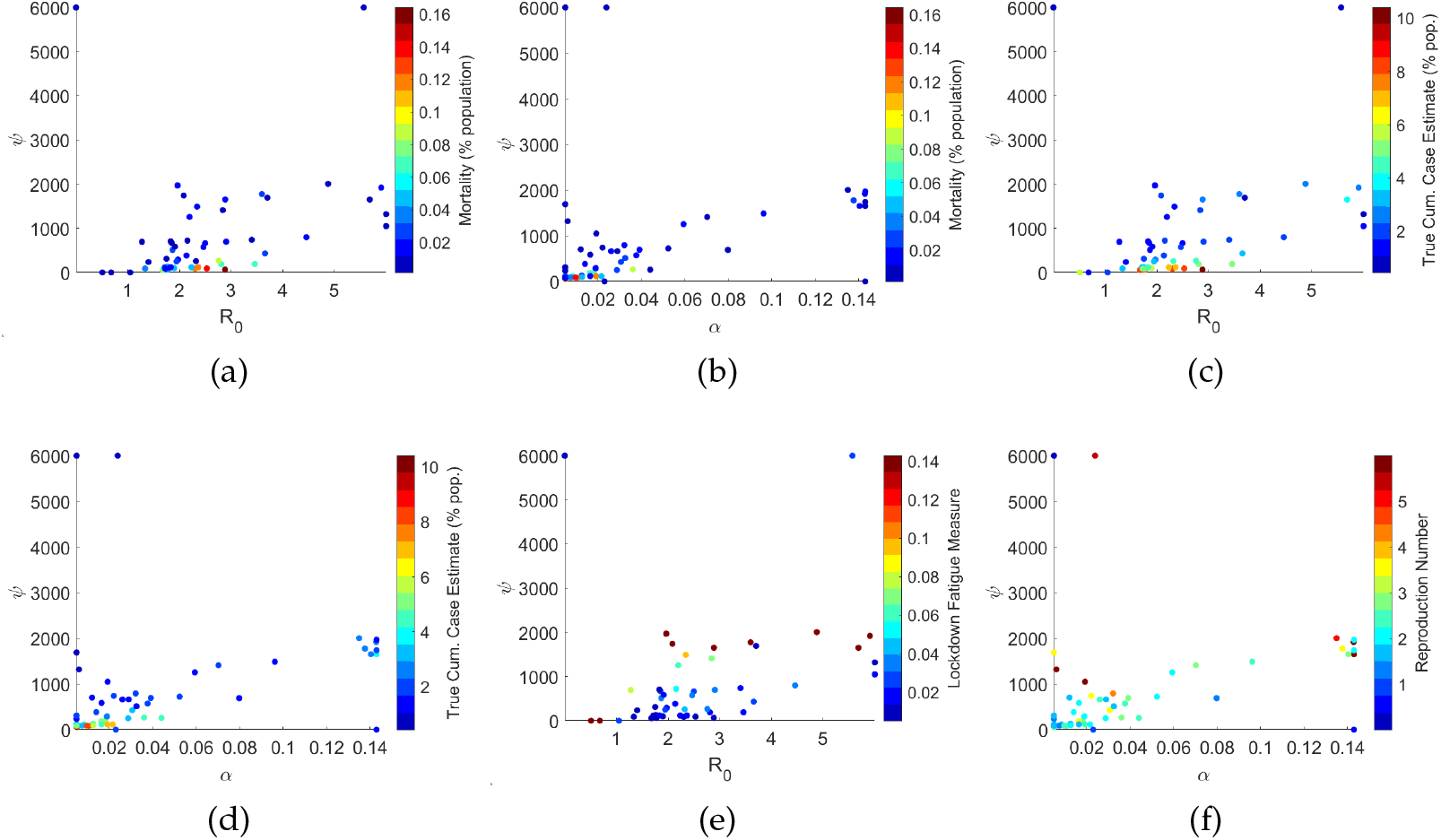
three-variable scatter plots for various model parameters from state and territory fits. *ψ* dominates the effect of all other model parameters. Of particular note is that high *ψ* values tend to be associated with high *α* values.

Furthermore we observe that *ψ* and estimated true case totals (shown in 5 (c)) as well as peak daily case totals ((d) of the same figure) follow an inverse proportionality relationship with respect to *ψ*. Indeed, as derived in [4], for the case *α* = 0 (indefinite quarantine period), the final cumulative infected, 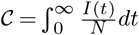 and peak infected, 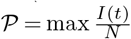, (as % of population) of model (2.1) satisfy

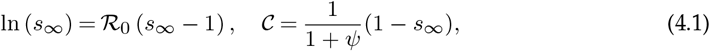

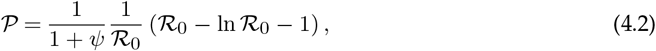

where 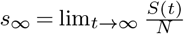 and *I*_0_ *≈* 0 (at start of outbreak). In each formula, the factor 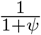 is multiplied by the corresponding classical relations for final and peak outbreak size. In view of our model fitting, the true cumulative and peak case estimates of each state fall roughly into an inverse proportionality with *ψ*, although states with high lockdown fatigue (*α* much larger than zero) stray from this pattern, as shown in Figure 6. In particular, by simply calibrating the inverse proportionality relation, 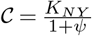, to the estimated parameters and cumulative cases of New York (which has small *ψ* and *α ≈* 0), we approximately fit the rest of the state values, with the analogous result for peak cases (see Fig. 6). The different values of ℛ_0_ and *α* in each state, however, shift the respective case numbers from the expected values predicted by inverse relation with *ψ* (Fig. 6). Thus while *ψ* has strongest impact, with rapidly escalating costs as the lockdown (rate) factor decreases below a critical value, ℛ_0_ and *α* still have an influence on outcome. We will examine these relationships further with counterfactual simulation studies.

Also of note is the suggested relationships between ℛ_0_ and *I*_0_ as well as ℛ_0_ and time to peak daily case incidence. Figure 5 panel a, which shows the aforementioned positive correlation between estimated *I*_0_ and date of first reported case (in terms of days after Jan. 21), and figure in the SI, which shows the negative correlation between ℛ_0_ and time to peak daily cases. This is further supported by the plots of time variable ℛ_*e*_ in Figure 2 panel (a) where it can be seen that in general states which have a later date for their first reported case tend to have higher reproduction number (for specific values see table 2). Other relationships of note are the positive relationship between IFR and cumulative case estimate as well as IFR and reported case estimate as well as the negative relationship between IFR and ratio of true to reported cases. This first relationship is likely explained by demographics and the law of large numbers: with higher cumulative case totals the likelihood both that hospital systems are overwhelmed and more individuals and clusters in high risk categories will become infected increases (for all fit IFR values see table 2). The last two relationships likely explain each other: as more cases and deaths were recorded greater effort was put into case reporting. And even in locales with successful outbreak responses such as Finland [11], or Hawaii in our own model, we note that the proportion of cases undetected in the early states of the outbreak is very high. For our United States fit we estimated an IFR of 0.009 (95% confidence interval .007 to .011), which is in line with the observed IFR for countries with very high levels of testing such as South Korea [16] and Finland [11].

Correlation analysis alone does not give the full picture. To further examine and differentiate the effects of *α, ψ* and ℛ_0_ on epidemic trajectories and suggest potential alternative strategies for management of subsequent waves of COVID-19 we carried out both counterfactual simulation studies and sensitivity analysis on these key model parameters. This was done using the fit model parameters for selected states which broadly speaking represent the range of outbreak responses which occurred in the United States. These are, in order of increasing cumulative outbreak size as of May 31 (see Figures 6,7,8):

i. Rapid Lockdown with Low Fatigue (RLLF) as represented by Hawaii
ii. Rapid Lockdown with High Fatigue (RLHF) as represented by Tennessee
iii. Intermediate Lockdown with Intermediate Fatigue (ILIF) as represented by Georgia
iv. Slow Lockdown with Low Fatigue (SLLF) as represented by New York

**Figure 4:**
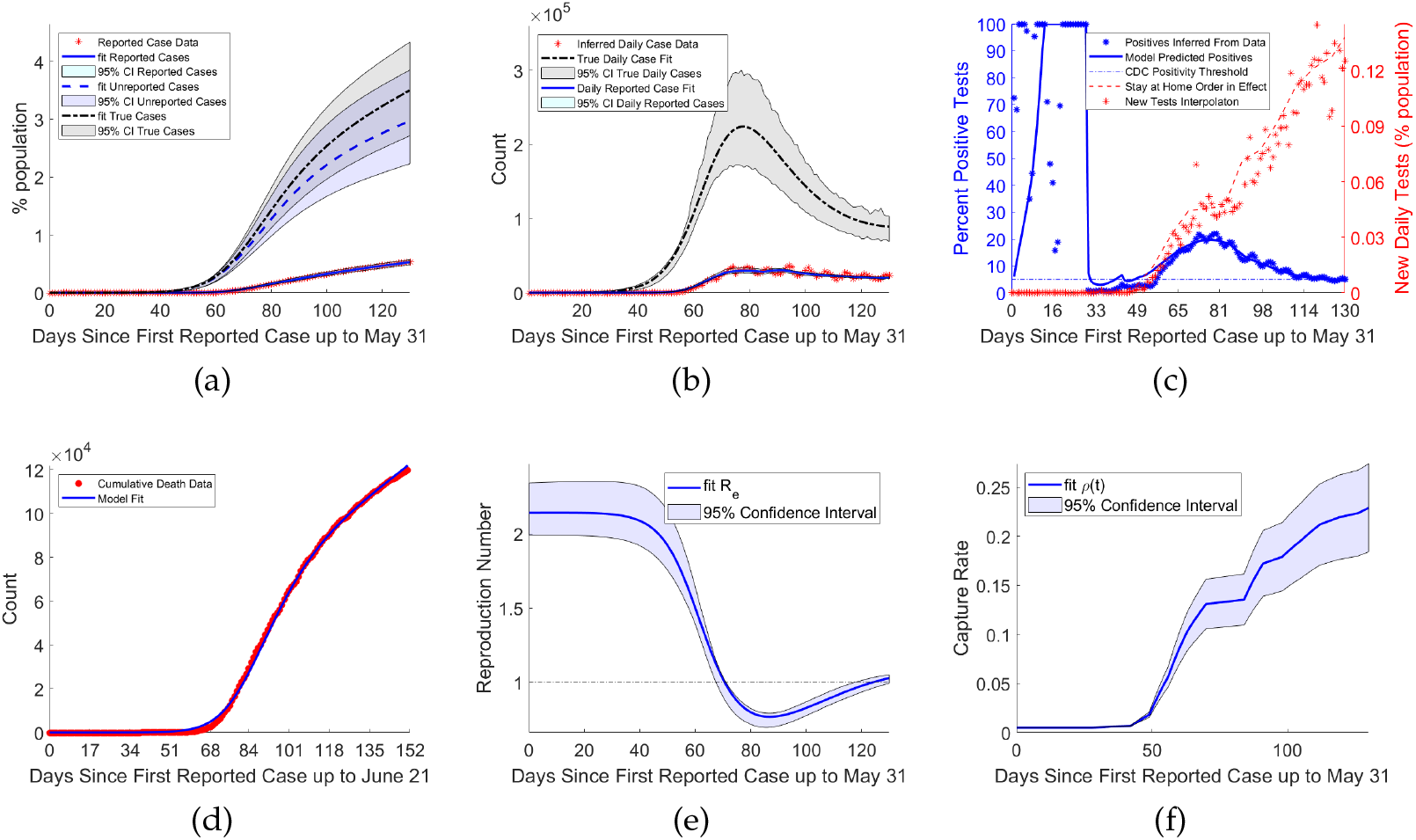
Model fit together with 95% confidence intervals for United States Fit

**Figure 5:**
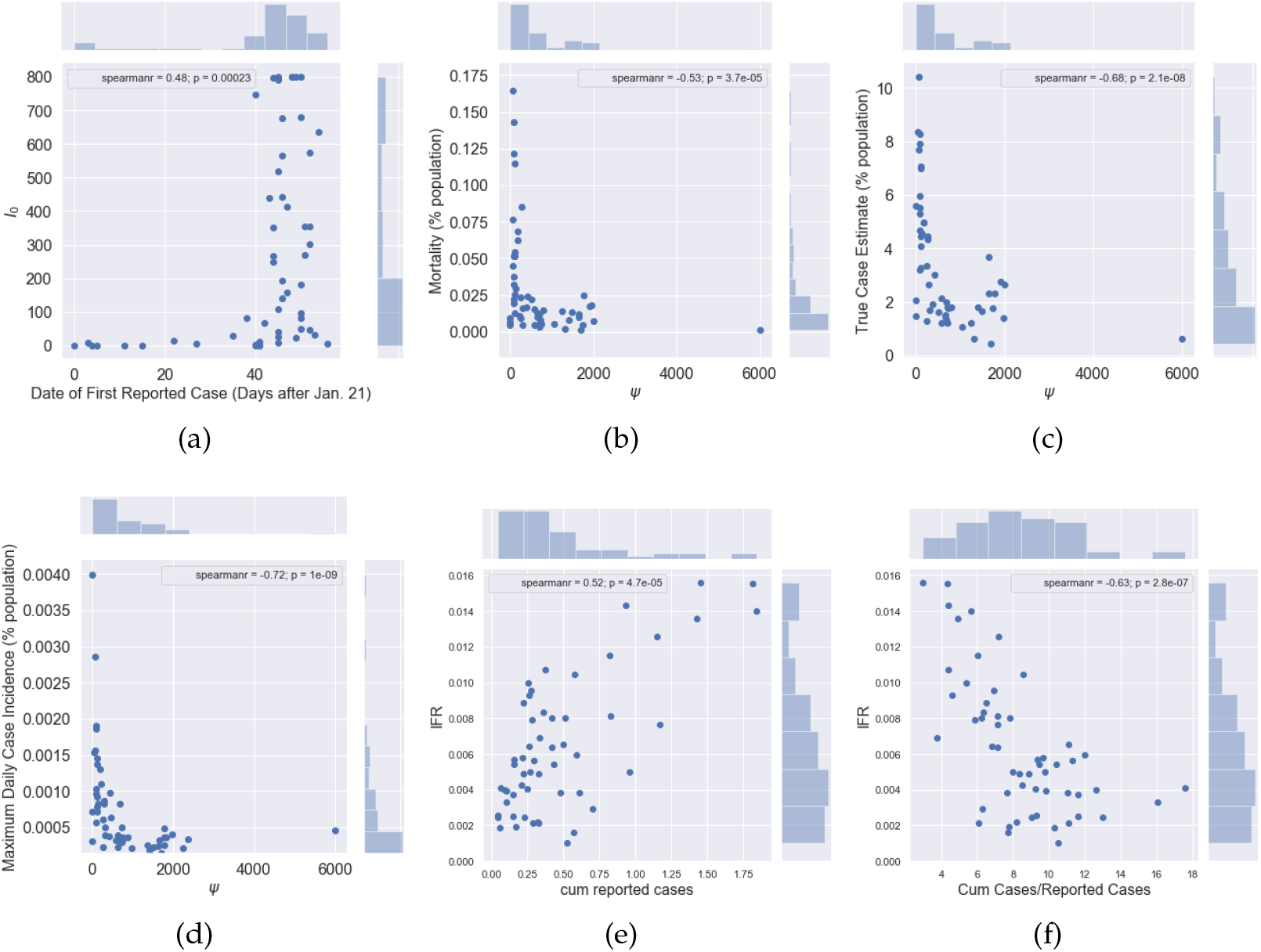
For Correlation Analysis The Northern Marianas Islands were not included because the fit ℛ_0_ value suggests all cases were imported (see table 2). (a) *I*_0_ vs Date of First Reported Case (days after Jan 21) (b) Mortality (% pop) vs *ψ* (c) True Case estimate (% pop) vs *ψ* (d) Peak daily cases (% pop) vs *ψ* (e) cumulative reported cases vs. IFR (f) Cum. Case Estimate/Reported cases vs IFR.

**Figure 6:**
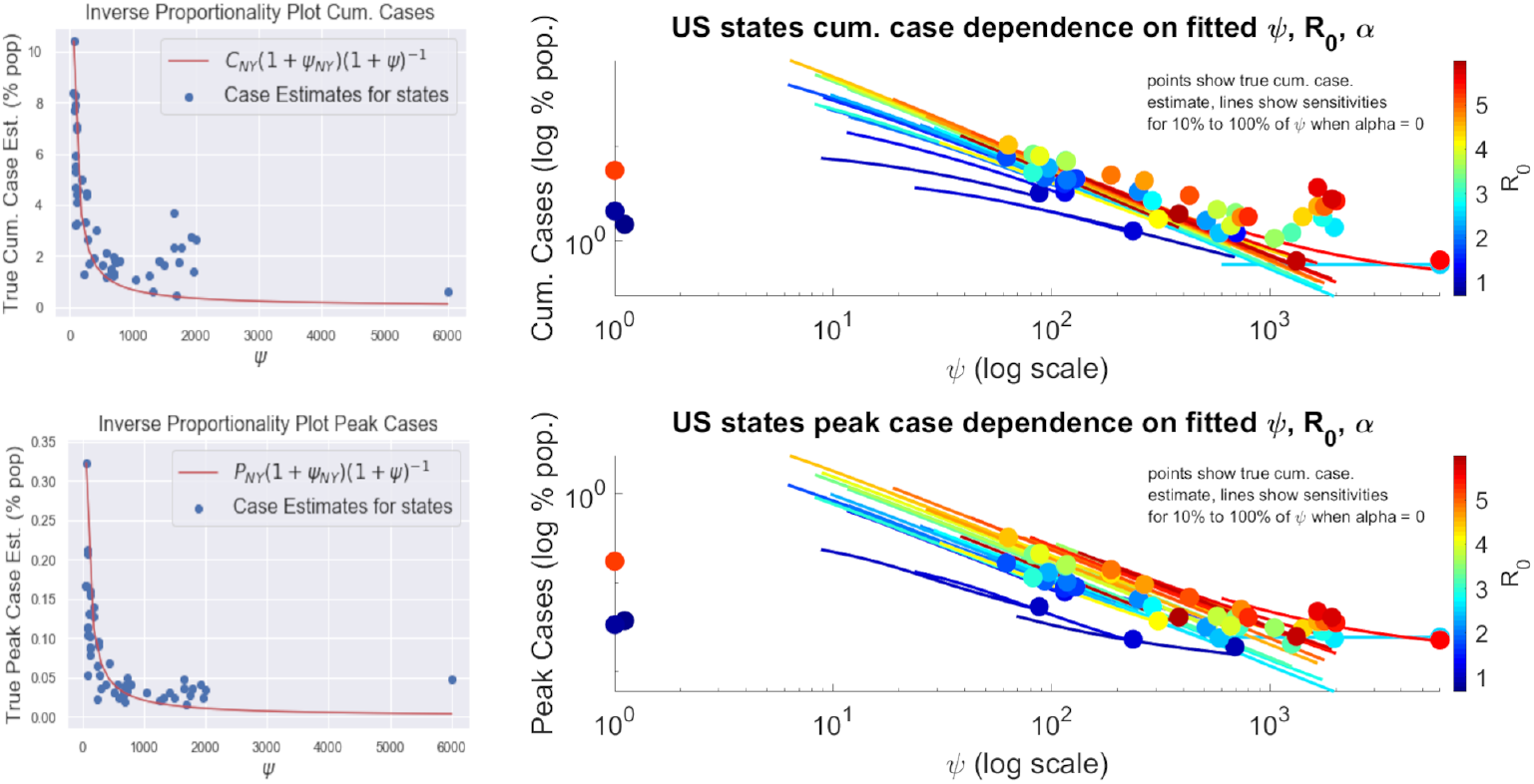
Both Cumulative cases and *ψ* (top right) and peak cases (bottom right) and *ψ* follow the inverse proportionality relationship described in [12]. The plotted lines indicate change in cumulative (peak) case estimate as *ψ* is varied from 10% it’s fit value up to the fit value itself. the color gradient of the lines from dark blue to dark red is indicative of increasing reproduction number ℛ_0_ This suggests that while *ψ* dominates these relationships, as our correlation analysis also indicates, *ℛ/* still has an impact on outcome with regard to both peak daily cases and cumulative case total. Top left shows the inverse proportionality relationship for the New York parameter set going from fit *ψ* to the maximum observed value of 6000. Bottom left is the same except for peak daily cases where the red lines follow equations (4.1) and (4.2) respectively

**Figure 7:**
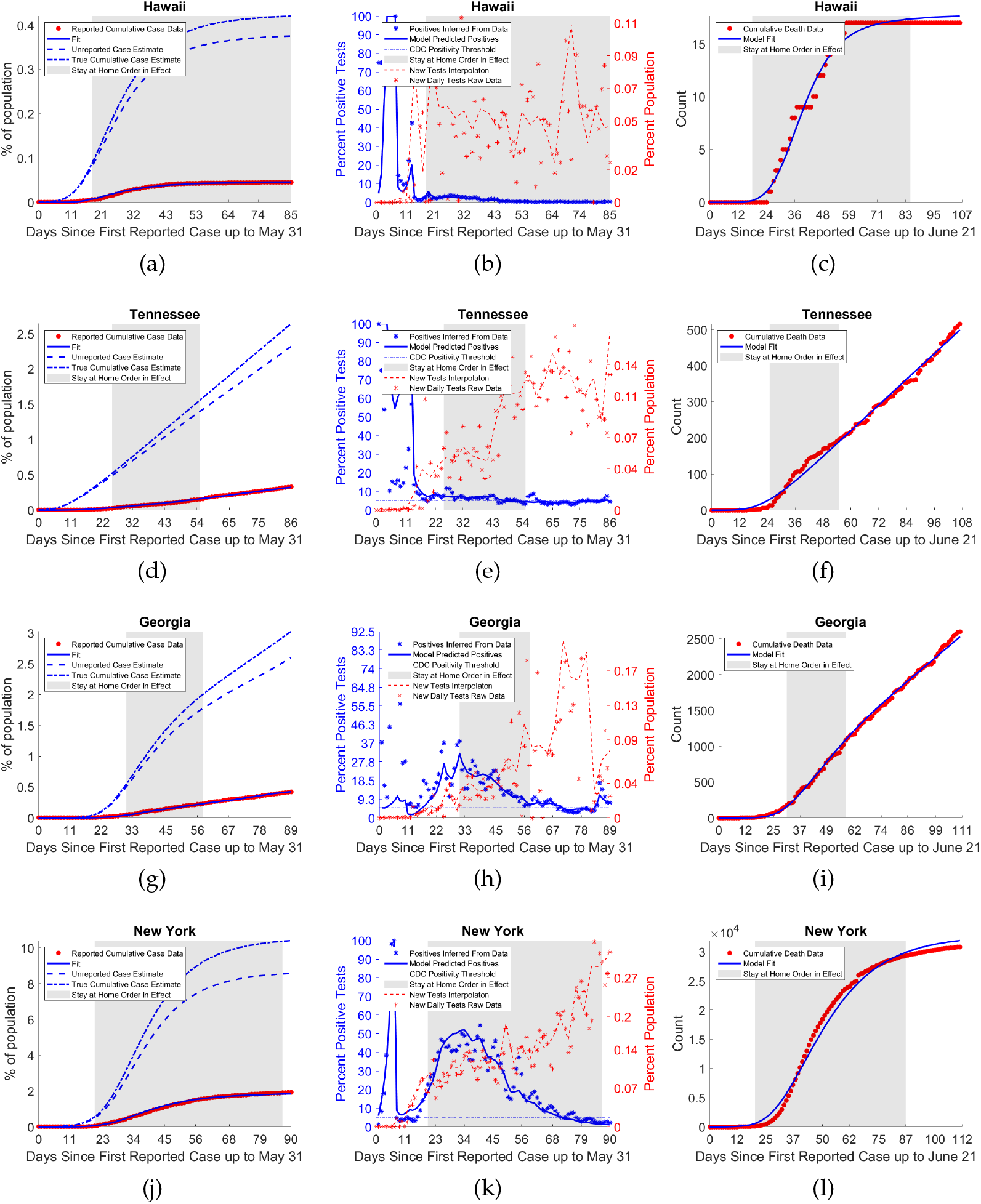
Representative examples of different relative value combinations of *ψ* and *α*, quality of strategy decreases from top to bottom row

**Figure 8:**
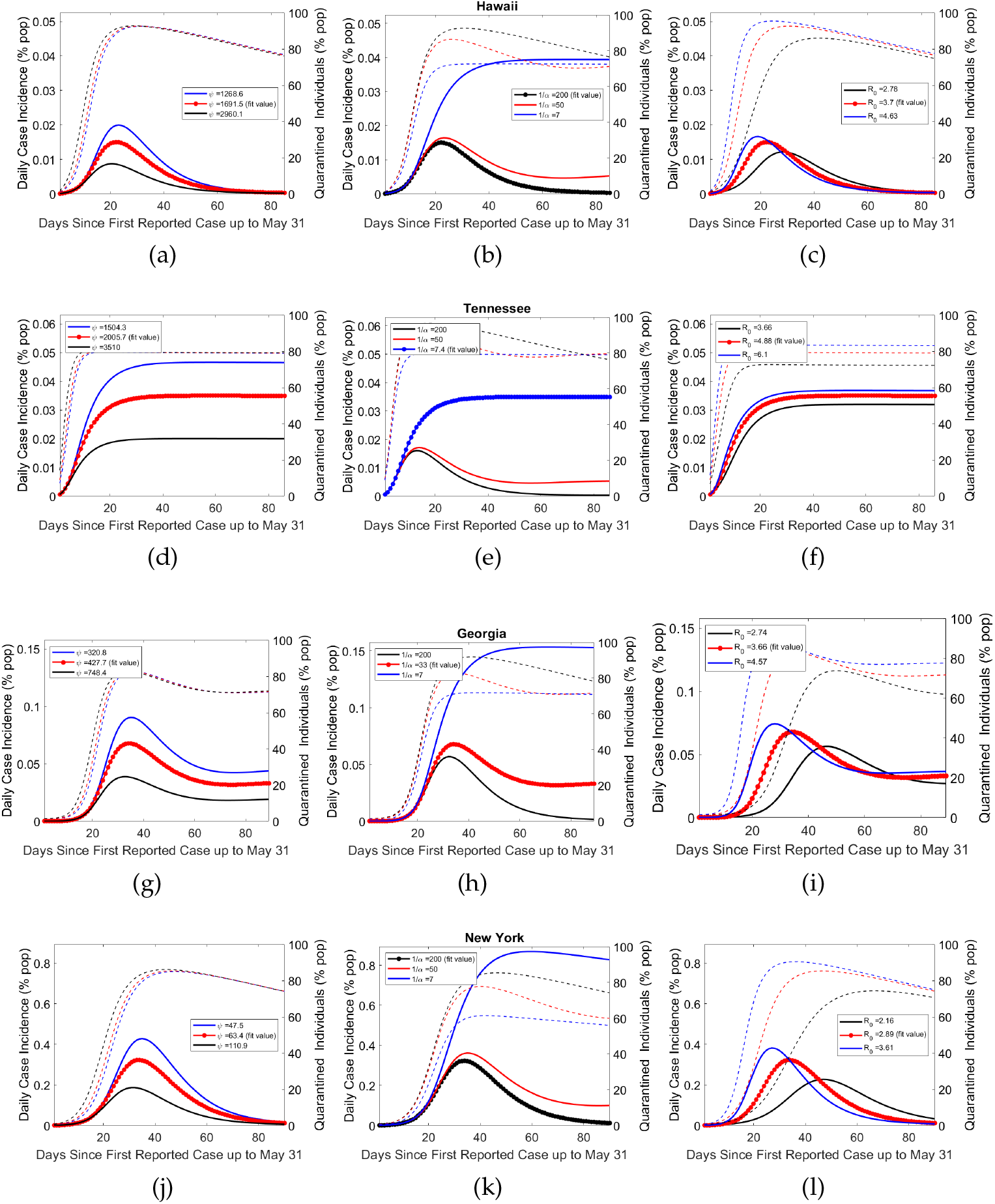
the differing effects of relative magnitudes and variation of *ψ, α*, and ℛ_0_ on epidemic trajectory as demonstrated by sample states shown in figure 6 and the whole United States Fit. For specific fit parameter values see table 2. Column 1 shows variation in *ψ*, column 2 in *α* and column 3 in ℛ_0_. Dashed lines indicate (self-)quarantined individuals over time corresponding to epidemic trajectory of the same color.

As can be seen in these figures the states all reported their first confirmed case within a week of one another, but had significantly different outcomes. Taken as percent of population Hawaii had both the lowest cumulative case total across all states and territories and one of the briefest outbreaks in terms of increasing daily case totals. All other considerations aside response RLLF represents the optimal strategy in terms of both peak and cumulative daily case totals.

Tennessee highlights the importance of lockdown fatigue. In the baseline fit epidemic trajectory, despite having a lockdown factor higher than that of Hawaii, Tennessee’s cumulative case totals are an more than 4 times as high as a result of significant linear growth in daily cases not far off the peak value. While this response will likely still keep hospitals from being overwhelmed provided sufficiently high *ψ* it both prolongs the outbreak, and leads to more cases than response RLLF.

Georgia represents a number of states for which both the fit *ψ* and *α* values falls into a middle ground between the maximal and minimal fit values across all states and territories. As can be seen in the fit trajectory and table 2 despite have a *ψ* value an order of magnitude lower than that of Tennessee and similar dates of first reported case, the two states have relatively similar cumulative case totals, suggesting that there may be a critical threshold for *α* which is examined further below.

New York is representative of the final observed response, slow lockdown with low lockdown fatigue. Despite having both the highest reported cumulative and estimated true case totals across all states and territories New York’s outbreak was essentially over at a time when many states were experiencing significant daily case totals despite similar dates of first reported case.

Our results suggest that the optimal response strategy in terms of case totals, and therefore also fatalities, remains rapid lockdown with low lockdown fatigue (response RLLF). Here the speed of lockdown is the critical factor as our model considers that individuals and/or government will eventually react to escalating cases, reaching similar numbers of self-quarantined (distinct for each state parameter set) in the counter-factual simulations of varying *ψ* with the difference being delayed lockdown resulting in order of magnitude more cases. While no substitute for the rapid lockdown strategy, the next best intervention may be sustained public social distancing and mask wearing, targeting transmission reduction rather than removing susceptibles all together, to reduce ℛ_0_. This has the added benefit of reducing the number of individuals that need to be quarantined, with those that are able to self quarantine doing so preferentially throughout the entire outbreak, but with at least half the quarantined population not returning to normalcy for a period of 30 days (see figure insert). Indeed, across all sample states, and regardless of model parameters, self-quarantine periods lasting longer than this threshold greatly reduce the daily case total on May 31, suggesting that if this is achieved other methods of managing the outbreak, such as contact tracing, could then be employed to manage subsequent cases.

Sufficiently high lockdown fatigue can lead to sustained linear growth in cumulative cases (or sustained steady state of daily cases) regardless of *ψ* value. Observe that despite Tennessee and Hawaii having fairly similar fit *ψ* values, both the daily case load on May 31 and the peak daily case incidence for Tennessee are an order of magnitude higher than those of Hawaii when *ψ* is reduced by 90% (see Figure 8). Comparatively this difference is closer to a factor of five in baseline simulations. Figure 9 suggests that this is attributable to their different *α* values, which fall on opposite ends of the considered range. Thus our model suggests that lockdown fatigue, *α*, primarily affects whether or not daily case incidence on May 31 is close to zero relative to peak cases, where lockdown factor (or speed), *ψ*, modulates the peak level. A similar result is suggested with respect to ℛ_0_ and peak daily case total as well as date of peak, importantly with high *α* significantly dampening the impact of lowered ℛ_0_ (see Figures 8,9). While caution must be exercised with short duration or high-turnover lockdowns (responsive to accumulating cases), governments and individuals are also reluctant to return to strict lockdown. Given this reality, while response RLLF has the best outcomes, it is important to consider the viability of alternative strategies which might be implemented for future outbreaks.

**Figure 9:**
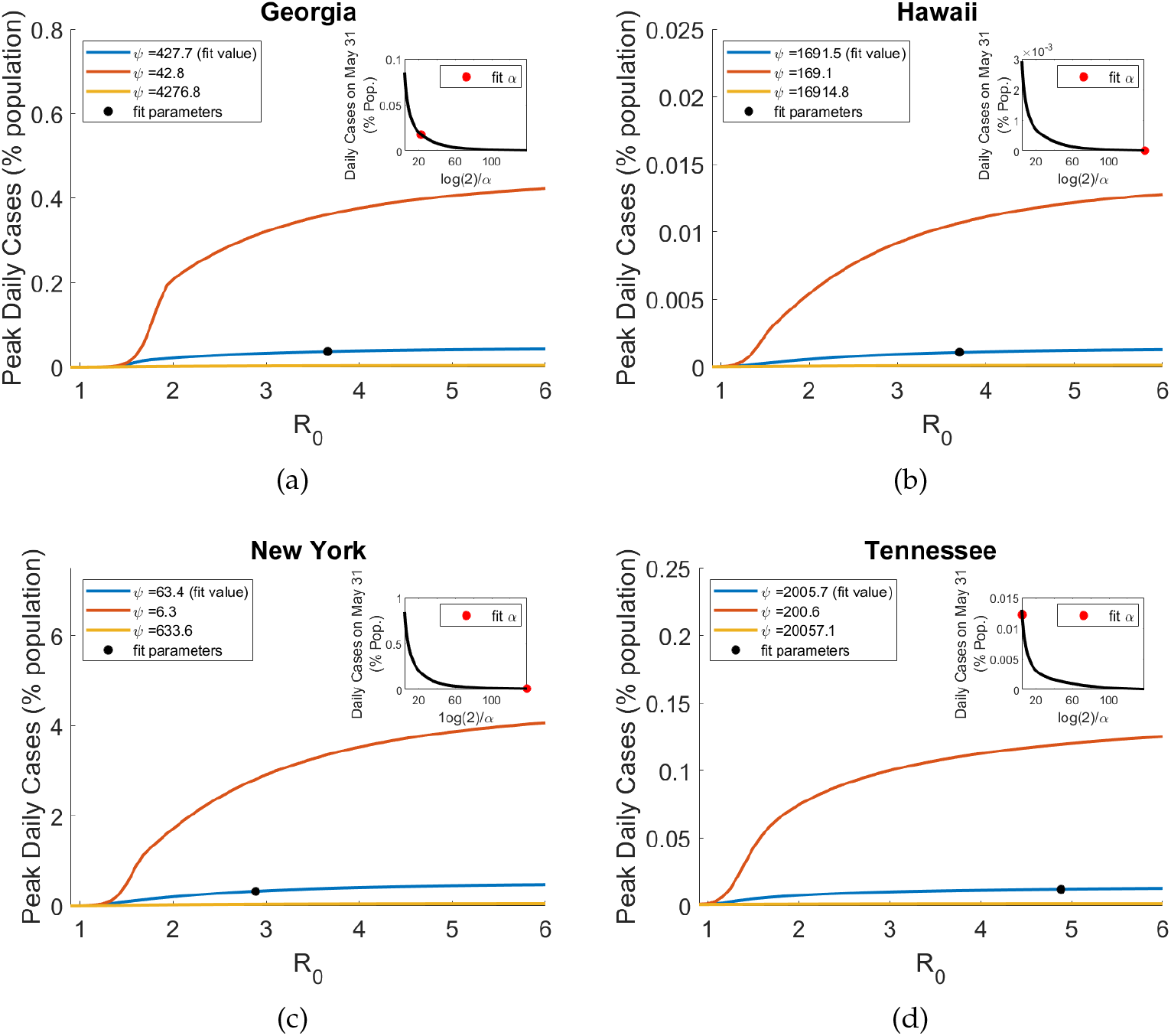
Peak daily cases as a function of ℛ_0_, magnitude of change is primarily determined by ψ as can be seen by the scales of the y-axes. *α* determines duration of outbreak, with critical half-life threshold of 30-60 days.

## 5. Discussion

In this study we fit a unified model to case, death, and testing data from all 50 states as well as Washington DC and four outlying territories in order to quantify impacts of a wide variety of lockdown entry/exit responses, which we correlate with true case estimates for insight into the dynamic COVID-19 outbreak control in the United States. Despite the heterogeneous nature of exhibited responses to the outbreak within each state, we were able to obtain quality fits for all considered states and territories. A crucial step in doing so was incorporating testing data, *τ* (*t*), by calibrating the ascertainment rate, *ρ*(*τ* (*t*)), as a saturating function of per-capita tests, which provided extra confidence in our fitting because daily percent positive tests were mimicked in each state. The inferred true cumulative case totals demonstrate high varying levels of under-reporting, with an estimated 6.62 cases per reported cases (cumulative) overall in the US up to May 31, 2020. Furthermore, our modeling framework allows us to infer Infection Fatality Ratio, found to be 0.9% for the US over time period. These are likely only estimates for (reported and unreported) symptomatic cases which do not account for a significant extent of asymptomatic individuals, since testing, reported case, and mortality data are all derived mostly from symptomatic infections.

Our model captured increasing case ascertainment rates through time (as testing increased), with a significant positive correlation to the amount of cases reported in a state, in contrast to a recent study in France showing a negative relationship between ascertainment ratio and reported cases as larger case counts hampered their substantial contact tracing program [11]. In the United States, comparatively little effort was put into contact tracing, and the particular geographical or political features of COVID-19 spread, along with higher case burden focusing community attention on the disease, may have contributed to the reverse correlation. Faced with a spreading outbreak, high numbers of undetected infection, and no nationwide control policy, the US adopted heterogeneous responses centered around some form of lockdown, which we characterize according to parameter fits of quarantine entry/exit rate for each state.

Our model fitting results suggest, broadly speaking, that the distinct responses can be divided into one of four categories (arranged in order of generally increasing cumulative case totals): Rapid Lockdown with Low Fatigue (RLLF), Rapid Lockdown with High Fatigue (RLHF), Intermediate Lockdown with Intermediate Fatigue (ILIF), and Slow Lockdown with Low Fatigue (SLLF). Using correlation analysis of the key parameters and outcomes over all US states, along with counterfactual simulation studies for representative states of these range of responses (Hawaii, Tennessee, Georgia, and New York), we translate the unified model fits into assessing the variable aspects of reactive lockdowns or self-quarantine (speed, scale, duration/fatigue) in combination with sustained interventions aimed at reducing ℛ_0_, such as public face mask wearing and social distancing, contact tracing.

Lockdown (rate) factor *ψ* played the largest role in outbreak severity, in particular both cumulative and peak cases roughly followed an inverse proportionality relationship with *ψ*, which was theoretically derived and applied to the epidemic in China [12] in the instance self-quarantine (lockdown) exit rate, *α*, is zero. Differences in ℛ_0_ also influenced outbreak shape, where its reduction mainly results in both a delayed peak in daily case totals and decreased size of peak daily case incidence, along with lessening the scale of lockdown needed to control the diseases. High lockdown fatigue (*α* significantly larger than zero) in some US states did impact their outbreaks though, the daily case load at the end of the time-frame considered (May 31) being large, with magnitude determined primarily by *ψ*, but also ℛ_0_. Importantly, independent of all other model parameters our model suggests a critical threshold of 30-60 days for half-life return to normalcy, that is for 50% of the quarantined population to exit lockdown (see Figure 9). With this threshold indicating the approximate point at which there is significant reduction in daily case load within approximately 90 days of first reported case, perhaps allowing for other less invasive control methods, such as contact tracing, to be implemented.

Our results provide insights for management of future outbreaks. The optimal strategy purely in terms of cumulative, and peak daily cases, and therefore also fatalities remains response RLLF (rapid lockdown with low fatigue). However, in order to balance economic/social concerns in addition to the purely epidemiological, we suggest a new response to an outbreak compared to those observed in the United states first wave; Rapid measured lockdown with intermediate fatigue. This strategy describes implementation of rapid reactive lockdown as soon as possible in conjunction with subsequent wave being detected, lasting at least 30 days before 50% return to normalcy, with the underlying ℛ_0_ and public adherence to proper social distancing and mask wearing determining the scale of closures during lockdown (where at-risk or sectors responsible for large-scale spread are preferentially included in quarantine). The quick response and critical duration of quarantined sectors will allow case numbers to be sufficiently reduced after the 30 day period for contact tracing to be feasible and in combination with broader measures aimed at lowering ℛ_0_ (e.g. face masks) can prevent any substantial subsequent wave until effective vaccines are widely taken by the population.

## Data Availability

All relevant data and code are available at:
https://github.com/jcmacdonald-codesData?tab=projects

## Supporting Information

SI Appendix

Figs. S1-S19

Table S1, S2

## Acknowledgment

JCM, CJB, and HG are supported by a U.S. National Science Foundation RAPID grant (DMS-2028728). HG was also supported by an NSF grant (DMS-1951759) and a grant from the Simons Foundation/SFARI(638193). CJB is partially supported by an NSF grant (DMS-1815095).

The authors would like to thank Fadoua Yahia (Department of Mathematics, University of Louisiana at Lafayette) for her contributions to an earlier version of this paper.

## Appendix

**Table 2:** key parameter values and inferred quantities. True Case estimate is as of May 31, 2020. Summary statistics exclude the N. Marianas Islands due to fit parameter set suggesting all cases were imported

| State | $\mathcal{R}_0$ | $\psi$ | $\alpha$ | IFR | $I_0$ | True Case (%) | True Case/Rep. Case |
| --- | --- | --- | --- | --- | --- | --- | --- |
| Alabama | 5.91 | 1919.8 | 0.143 | 0.005 | 353.5 | 2.77 | 8.38 |
| Alaska | 5.57 | 6000.0 | 0.024 | 0.002 | 353.5 | 0.62 | 10.29 |
| Arizona | 1.96 | 1968.3 | 0.143 | 0.010 | 1.2 | 1.38 | 5.38 |
| Arkansas | 1.05 | 1.0 | 0.023 | 0.002 | 800.0 | 2.06 | 9.04 |
| California | 2.20 | 1254.9 | 0.059 | 0.009 | 1.0 | 1.21 | 4.59 |
| Colorado | 1.76 | 129.0 | 0.013 | 0.005 | 798.2 | 4.56 | 10.42 |
| Connecticut | 2.30 | 82.5 | 0.011 | 0.013 | 411.6 | 8.28 | 7.21 |
| Delaware | 1.78 | 62.1 | 0.005 | 0.005 | 181.3 | 7.68 | 7.98 |
| Dist. of Columbia | 1.68 | 55.1 | 0.005 | 0.008 | 192.3 | 8.37 | 7.14 |
| Florida | 4.46 | 795.0 | 0.032 | 0.006 | 1.0 | 1.79 | 6.84 |
| Georgia | 3.66 | 427.7 | 0.031 | 0.006 | 10.1 | 3.02 | 7.14 |
| Guam | 0.51 | 1.0 | 0.143 | 0.001 | 634.7 | 5.58 | 10.54 |
| Hawaii | 3.70 | 1691.5 | 0.005 | 0.003 | 9.5 | 0.42 | 9.30 |
| Idaho | 3.40 | 738.2 | 0.022 | 0.002 | 573.6 | 1.77 | 11.65 |
| Illinois | 1.75 | 114.0 | 0.008 | 0.014 | 8.9 | 4.08 | 4.38 |
| Indiana | 1.75 | 90.0 | 0.007 | 0.007 | 792.5 | 5.53 | 11.08 |
| Iowa | 1.78 | 93.0 | 0.005 | 0.004 | 157.8 | 4.68 | 7.68 |
| Kansas | 5.68 | 1649.7 | 0.143 | 0.002 | 677.6 | 3.66 | 11.08 |
| Kentucky | 2.50 | 662.0 | 0.026 | 0.009 | 27.6 | 1.44 | 6.52 |
| Louisiana | 2.81 | 187.1 | 0.018 | 0.012 | 800.0 | 4.97 | 6.05 |
| Maine | 2.84 | 1409.7 | 0.070 | 0.004 | 270.1 | 1.79 | 11.63 |
| Maryland | 1.90 | 96.8 | 0.013 | 0.008 | 266.3 | 5.95 | 7.16 |
| Massachusetts | 2.36 | 117.1 | 0.019 | 0.014 | 1.1 | 6.98 | 4.90 |
| Michigan | 3.46 | 187.5 | 0.016 | 0.010 | 800.0 | 4.97 | 8.58 |
| Minnesota | 1.72 | 115.1 | 0.005 | 0.008 | 108.6 | 3.26 | 7.81 |
| Mississippi | 1.34 | 87.7 | 0.008 | 0.008 | 680.6 | 3.19 | 6.26 |
| Missouri | 2.47 | 572.4 | 0.038 | 0.006 | 140.1 | 2.14 | 9.68 |
| Montana | 6.00 | 1318.8 | 0.006 | 0.002 | 47.5 | 0.61 | 13.03 |
| Nebraska | 1.83 | 117.6 | 0.016 | 0.003 | 6.0 | 4.44 | 6.32 |
| Nevada | 1.97 | 289.2 | 0.019 | 0.005 | 250.3 | 2.65 | 9.77 |
| New Hampshire | 3.60 | 1775.0 | 0.138 | 0.008 | 1.0 | 2.30 | 6.33 |
| New Jersey | 2.53 | 88.3 | 0.010 | 0.016 | 440.3 | 7.92 | 4.36 |
| New Mexico | 1.87 | 510.1 | 0.033 | 0.011 | 81.0 | 1.63 | 4.38 |
| New York | 2.89 | 63.4 | 0.005 | 0.014 | 746.1 | 10.41 | 5.65 |
| North Carolina | 2.89 | 1652.0 | 0.140 | 0.004 | 67.4 | 2.31 | 9.24 |
| North Dakota | 1.40 | 235.8 | 0.005 | 0.007 | 49.9 | 1.27 | 3.75 |
| N. Marianas | 9.89e-12 | 6000.0 | 0.005 | 0.010 | 39.8 | 0.56 | 14.89 |
| Ohio | 1.94 | 249.9 | 0.029 | 0.006 | 800.0 | 3.34 | 11.32 |
| Oklahoma | 1.85 | 663.6 | 0.029 | 0.005 | 517.2 | 1.49 | 9.46 |
| Oregon | 1.92 | 585.5 | 0.017 | 0.004 | 82.2 | 1.20 | 12.63 |
| Pennsylvania | 2.23 | 116.2 | 0.022 | 0.006 | 800.0 | 7.07 | 11.98 |
| Puerto Rico | 1.75 | 307.5 | 0.005 | 0.003 | 300.2 | 1.69 | 16.09 |
| Rhode Island | 2.76 | 265.4 | 0.036 | 0.016 | 1.6 | 4.34 | 2.99 |
| South Carolina | 2.91 | 695.2 | 0.039 | 0.005 | 42.1 | 1.97 | 8.86 |
| South Dakota | 2.32 | 257.3 | 0.044 | 0.002 | 22.9 | 4.45 | 7.74 |
| Tennessee | 4.88 | 2005.7 | 0.135 | 0.002 | 352.1 | 2.64 | 8.17 |
| Texas | 2.15 | 721.3 | 0.052 | 0.004 | 14.7 | 1.80 | 8.54 |
| Utah | 2.08 | 1738.7 | 0.143 | 0.002 | 30.4 | 1.76 | 6.11 |
| Vermont | 0.68 | 1.1 | 0.143 | 0.006 | 442.7 | 1.48 | 9.34 |
| Virgin Islands | 1.83 | 702.4 | 0.012 | 0.004 | 33.3 | 1.21 | 17.58 |
| Virginia | 1.73 | 82.0 | 0.005 | 0.004 | 566.5 | 5.28 | 11.05 |
| Washington | 2.14 | 382.5 | 0.014 | 0.010 | 1.2 | 1.92 | 6.94 |
| West Virginia | 6.00 | 1048.5 | 0.019 | 0.004 | 5.0 | 1.05 | 9.83 |
| Wisconsin | 2.34 | 1487.4 | 0.096 | 0.008 | 1.0 | 1.67 | 5.86 |
| Wyoming | 1.27 | 691.0 | 0.080 | 0.002 | 95.9 | 1.32 | 7.78 |
| United States | 2.15 | 258.9 | 0.024 | 0.009 | 50.4 | 3.51 | 6.62 |
| mean | 2.593 | 714.06 | 0.0431 | 0.0064 | 275.02 | 3.359 | 8.404 |
| median | 2.176 | 344.99 | 0.0219 | 0.0055 | 148.95 | 2.474 | 8.079 |
| Std. Dev. | 1.330 | 962.41 | 0.0482 | 0.0038 | 293.75 | 2.362 | 2.939 |

**Table 3:** Approximate 95% Confidence Intervals, United States fit. Quantity is said to be practically identifiable if its Average Relative Error (ARE) is less than the noise level of 60%.

| Quantity | 95% CI | mean | median | ARE (%) |
| --- | --- | --- | --- | --- |
| $\mathcal{R}_0$ | (2.00, 2.38) | 2.19 | 2.04 | 4.53 |
| $\psi$ | (206.8, 348.5) | 289.0 | 294.0 | 15.60 |
| $\alpha$ | (0.020, 0.027) | 0.024 | 0.024 | 5.92 |
| IFR | (0.007, 0.011) | 0.009 | 0.010 | 15.09 |
| $I_0$ | (12.6, 134.0) | 48.0 | 41.2 | 55.40 |
| Cum. Case (%) | (2.72, 4.43) | 3.22 | 3.06 | 3.45 |
| True Cases/Rep. Cases | (5.54, 8.20) | 6.11 | 5.60 | 21.98 |

