## Supplementary Information for "Modeling COVID-19 outbreaks in United States with distinct testing, lockdown speed and fatigue rates"

### 1. Fitting Plots for All States and Territories

Plots of state and territory fits below grouped by region of the United States as determined by the Census Bureau and ordered from north to south within each region.

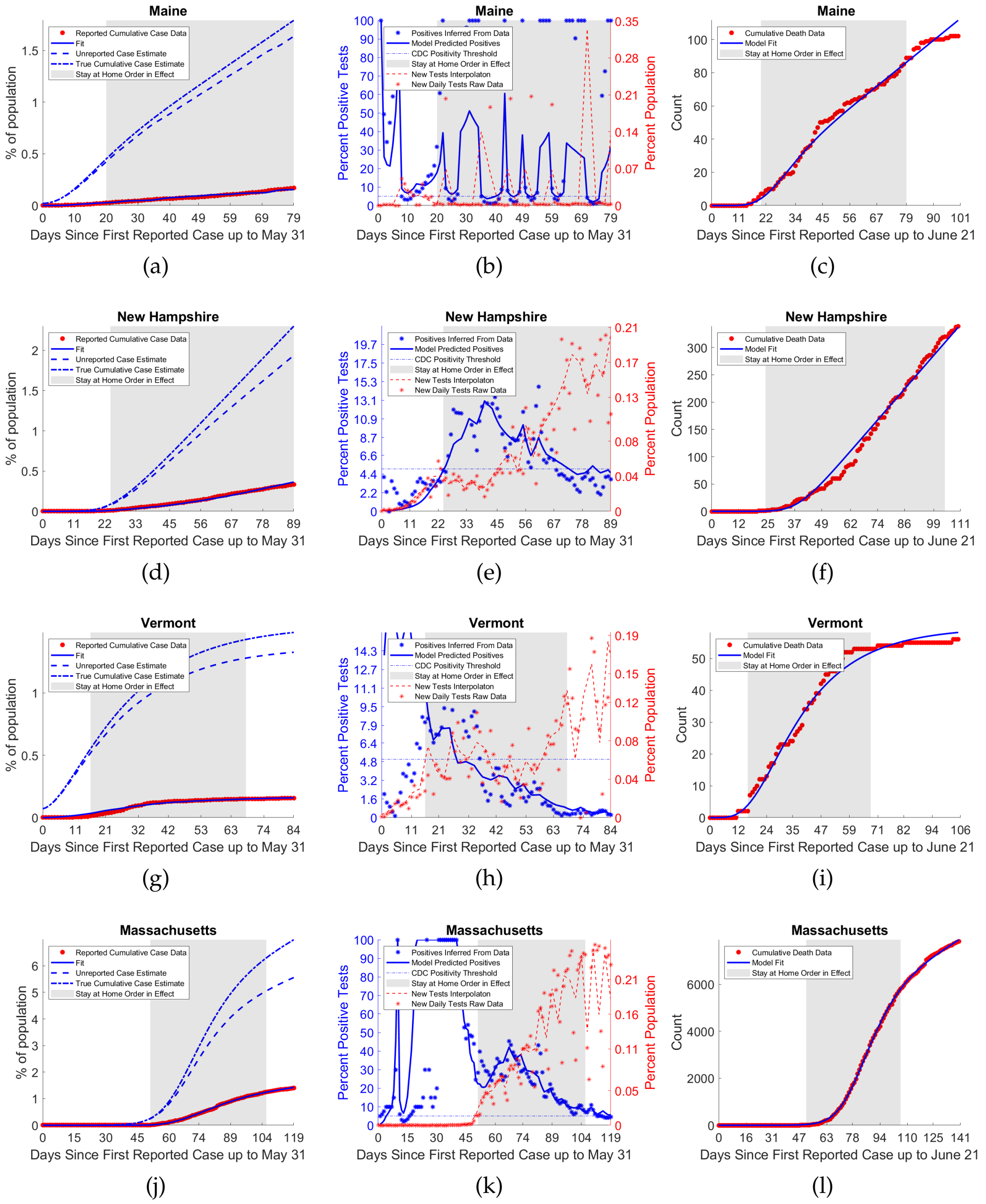


Figure S1: Fits for states in the Northeast. States and territories in this region include Maine, New Hampshire, Vermont, Massachusetts, Connecticut, Rhode Island, New Jersey, New York, and Pennsylvania. Figure 1 of 2. See also fig. S2

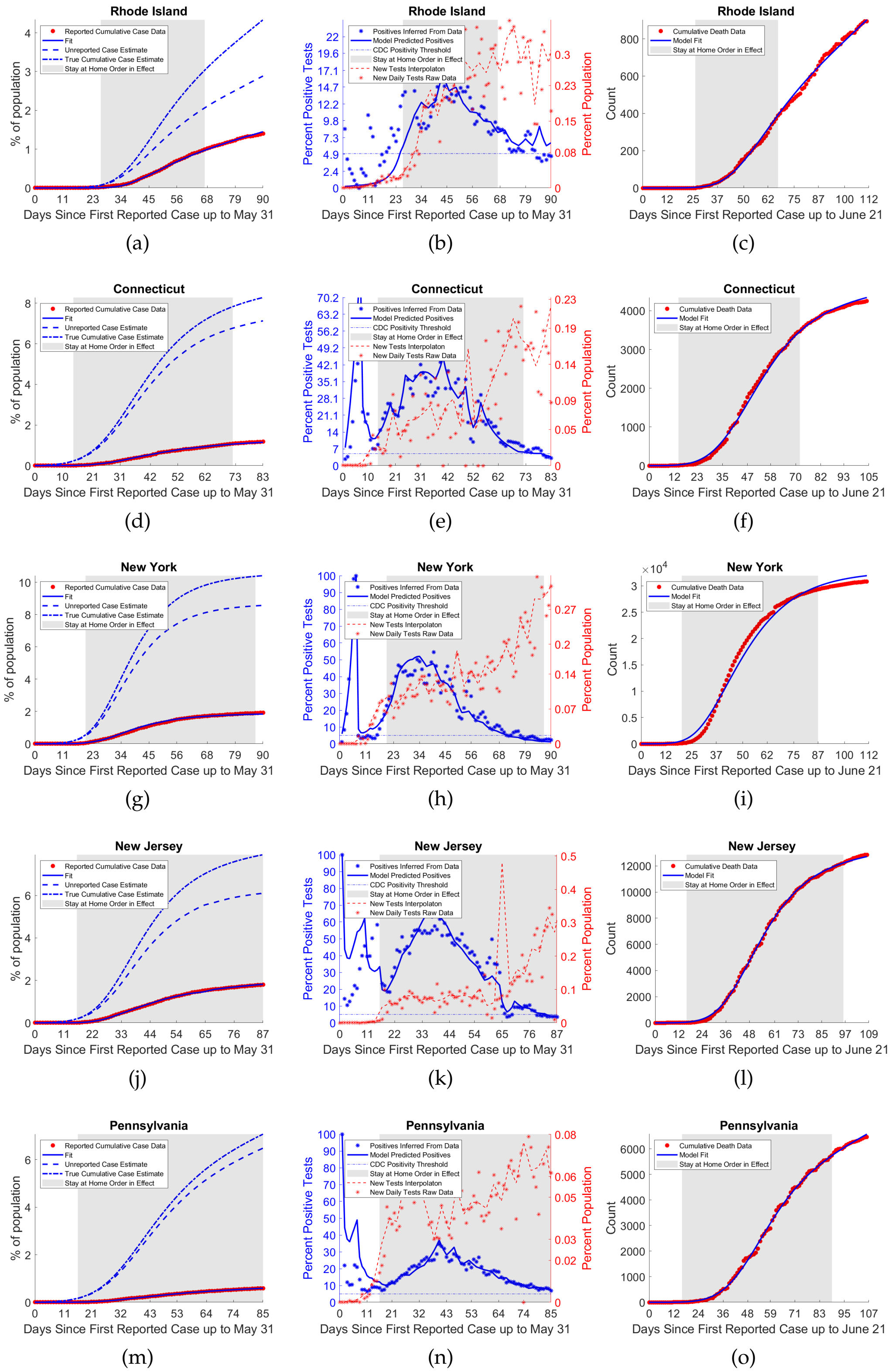


Figure S2: Fits for states in the Northeast. States and territories in this region include Maine, New Hampshire, Vermont, Massachusetts, Connecticut, Rhode Island, New Jersey, New York, and Pennsylvania. Figure 2 of 2.See also fig. S1.

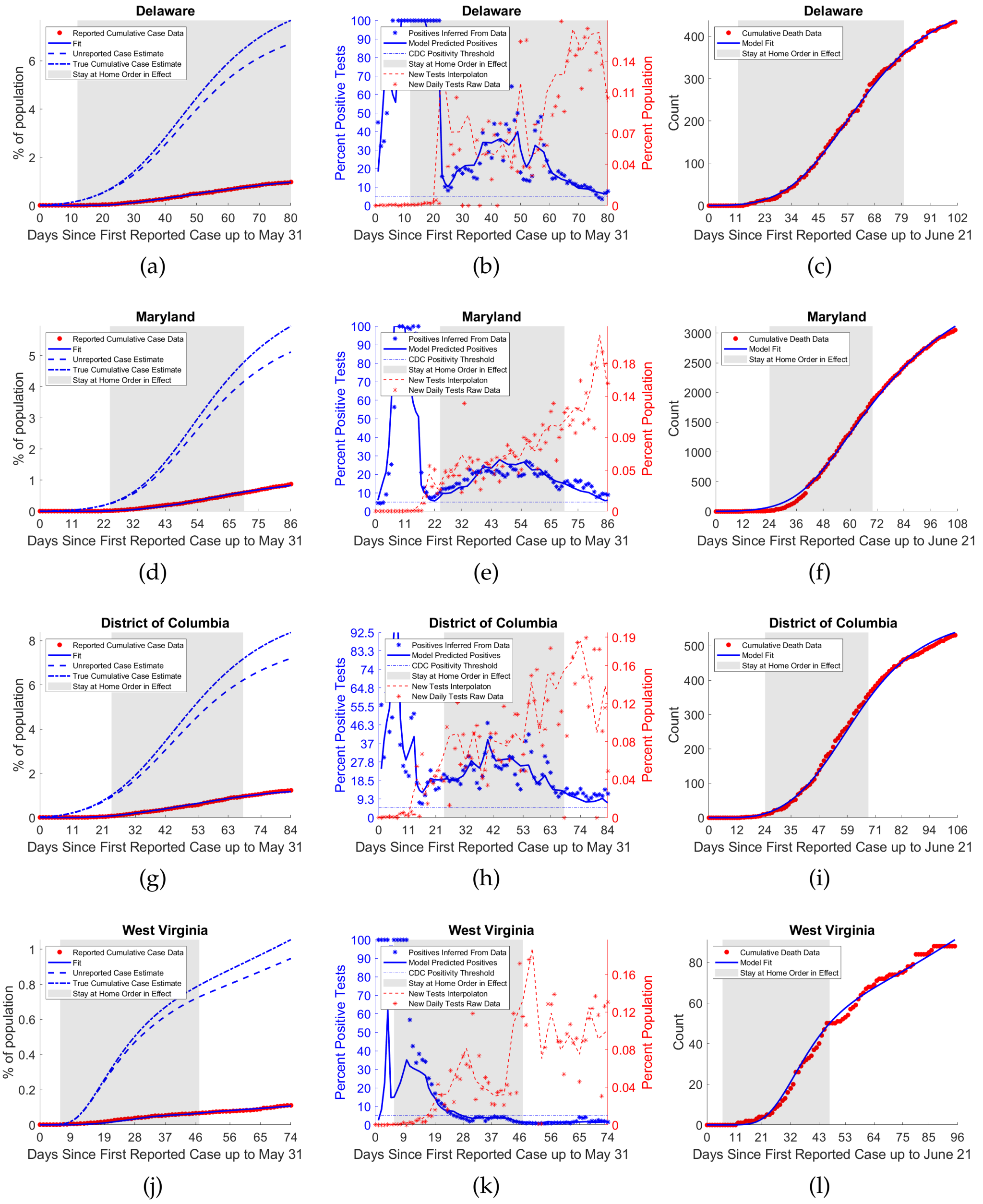


Figure S3: Fits for states and territories in the South Atlantic. States and territories in this region include Maryland, Delaware, West Virginia, Virginia, Georgia, North Carolina, South Carolina, Florida, the District of Columbia, the US Virgin Islands, and Puerto Rico. Figure 1 of 3. See also figs. S4,S5

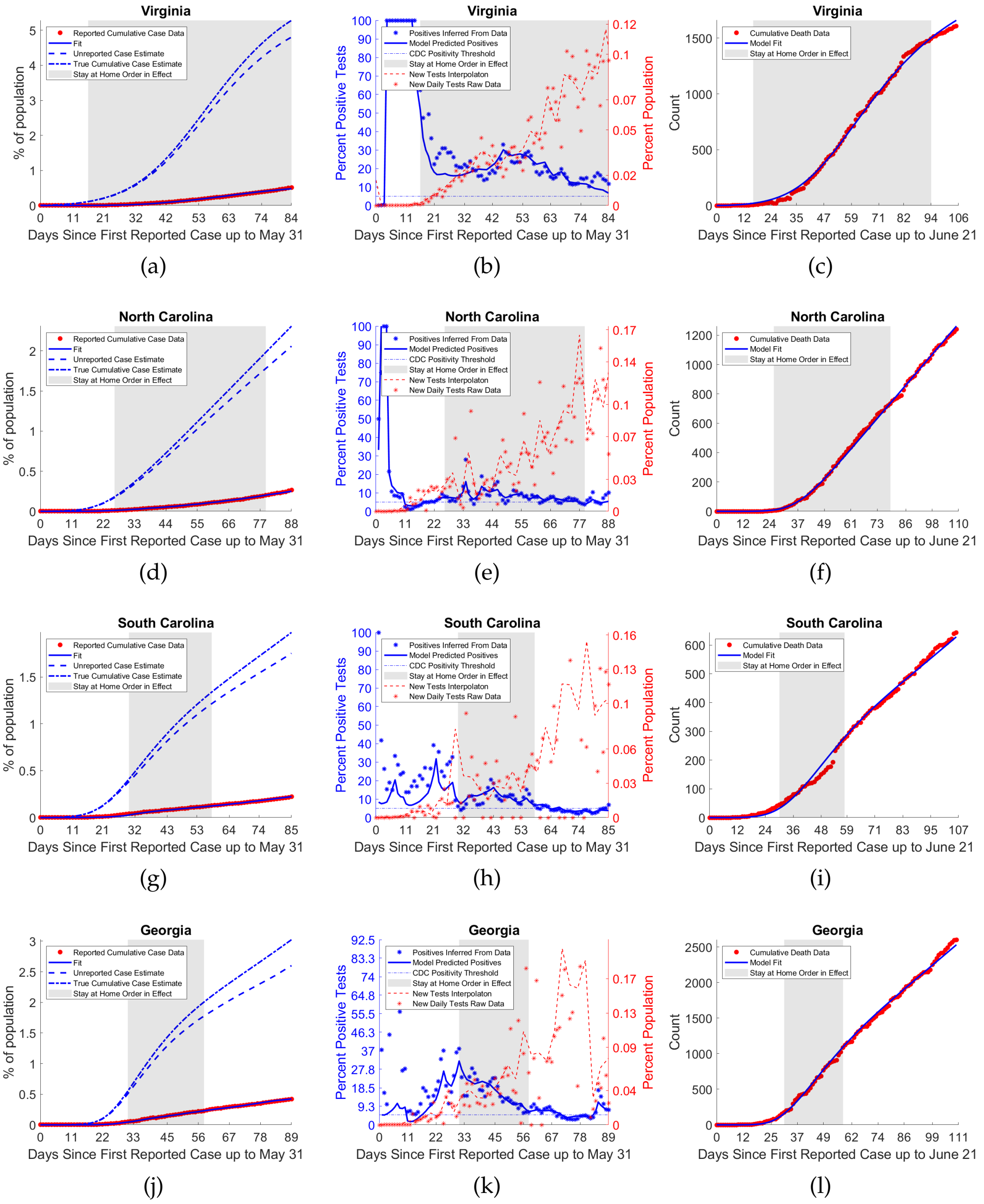


Figure S4: Fits for states and territories in the South Atlantic. States and territories in this region include Maryland, Delaware, West Virginia, Virginia, Georgia, North Carolina, South Carolina, Florida, the District of Columbia, the US Virgin Islands, and Puerto Rico. Figure 2 of 3. See also figs. S3,S5

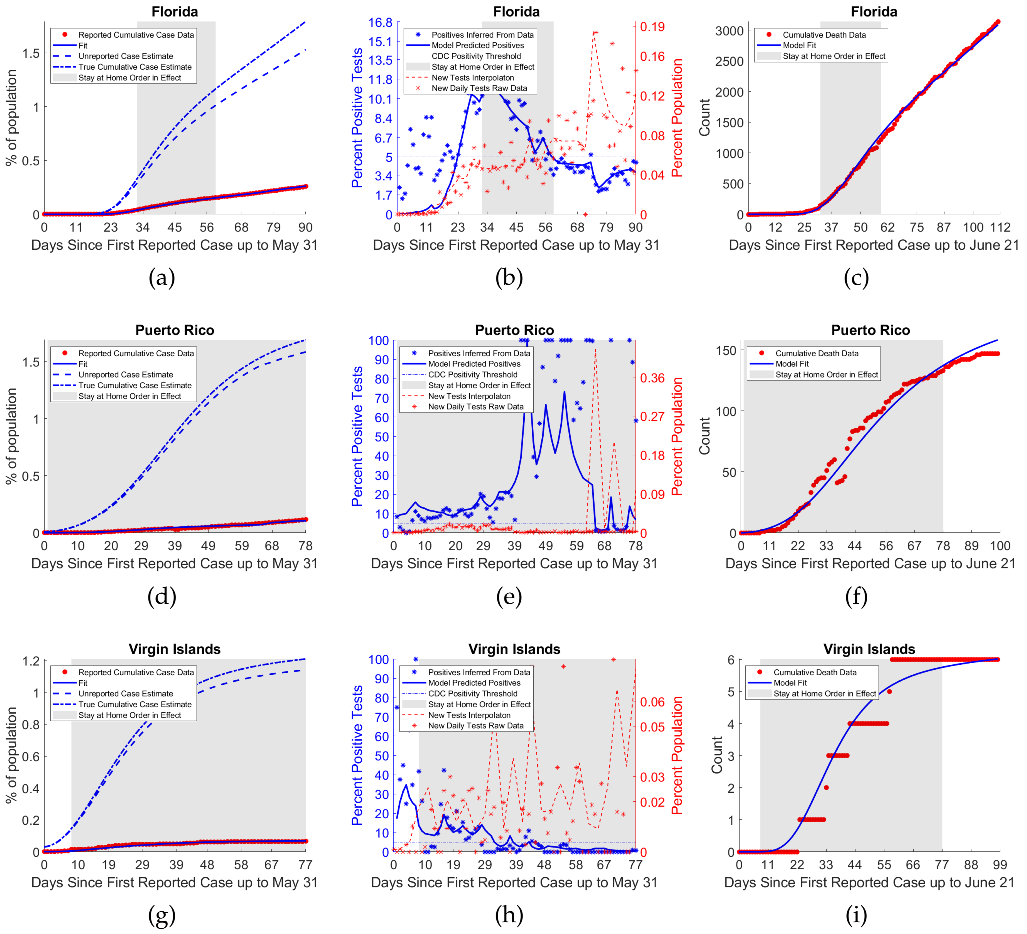


Figure S5: Fits for states and territories in the South Atlantic. States and territories in this region include Maryland, Delaware, West Virginia, Virginia, Georgia, North Carolina, South Carolina, Florida, the District of Columbia, the US Virgin Islands, and Puerto Rico. Figure 3 of 3. See also figs. S3,S4

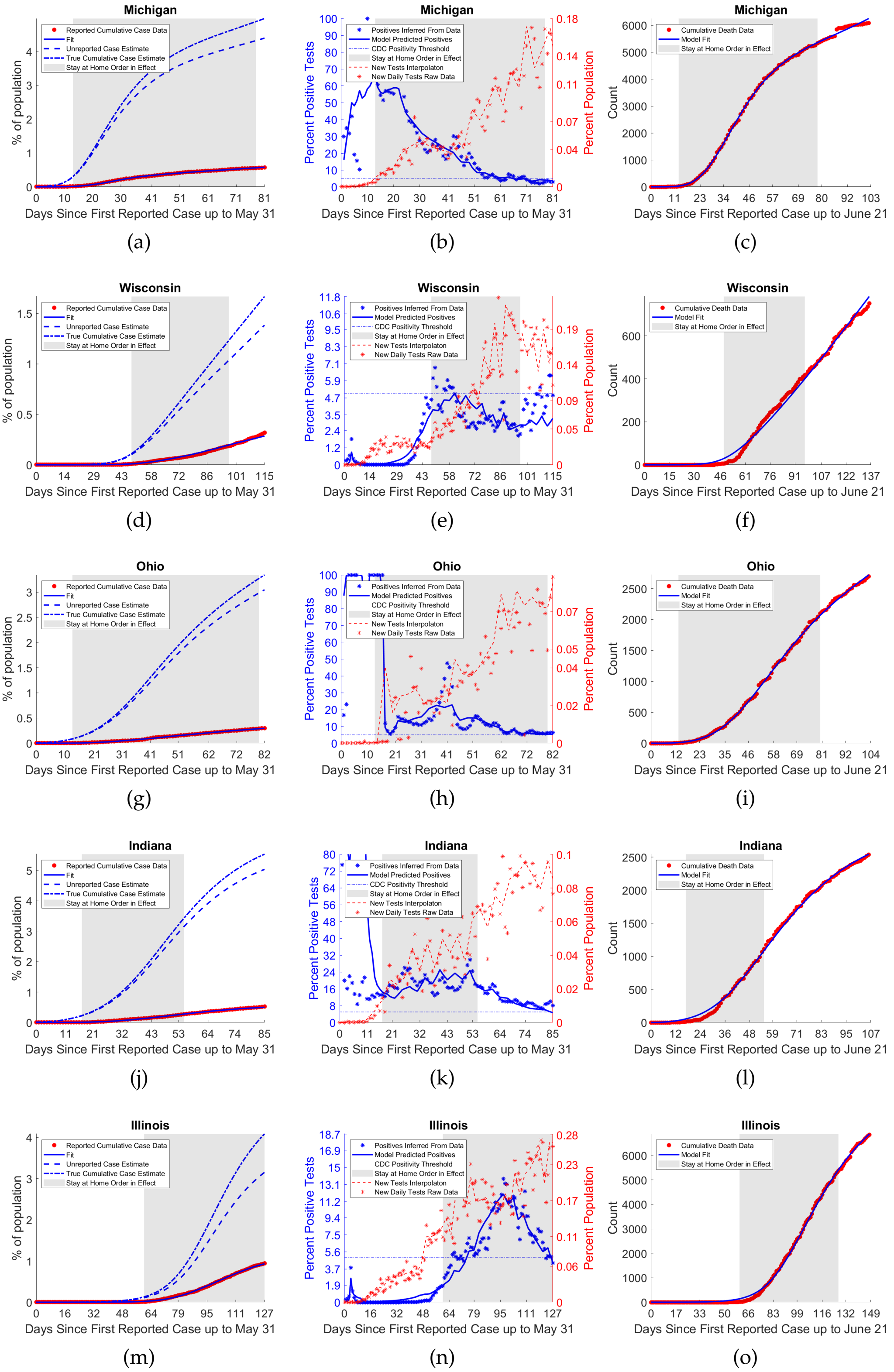


Figure S6: Fits for states and territories in the Midwest. States and territories in this region include Michigan, Wisconsin, Ohio, Indiana, Illinois, Minnesota, North Dakota, South Dakota, Iowa, Nebraska, Kansas, and Missouri. Figure 1 of 3. See also figs. S12,S8

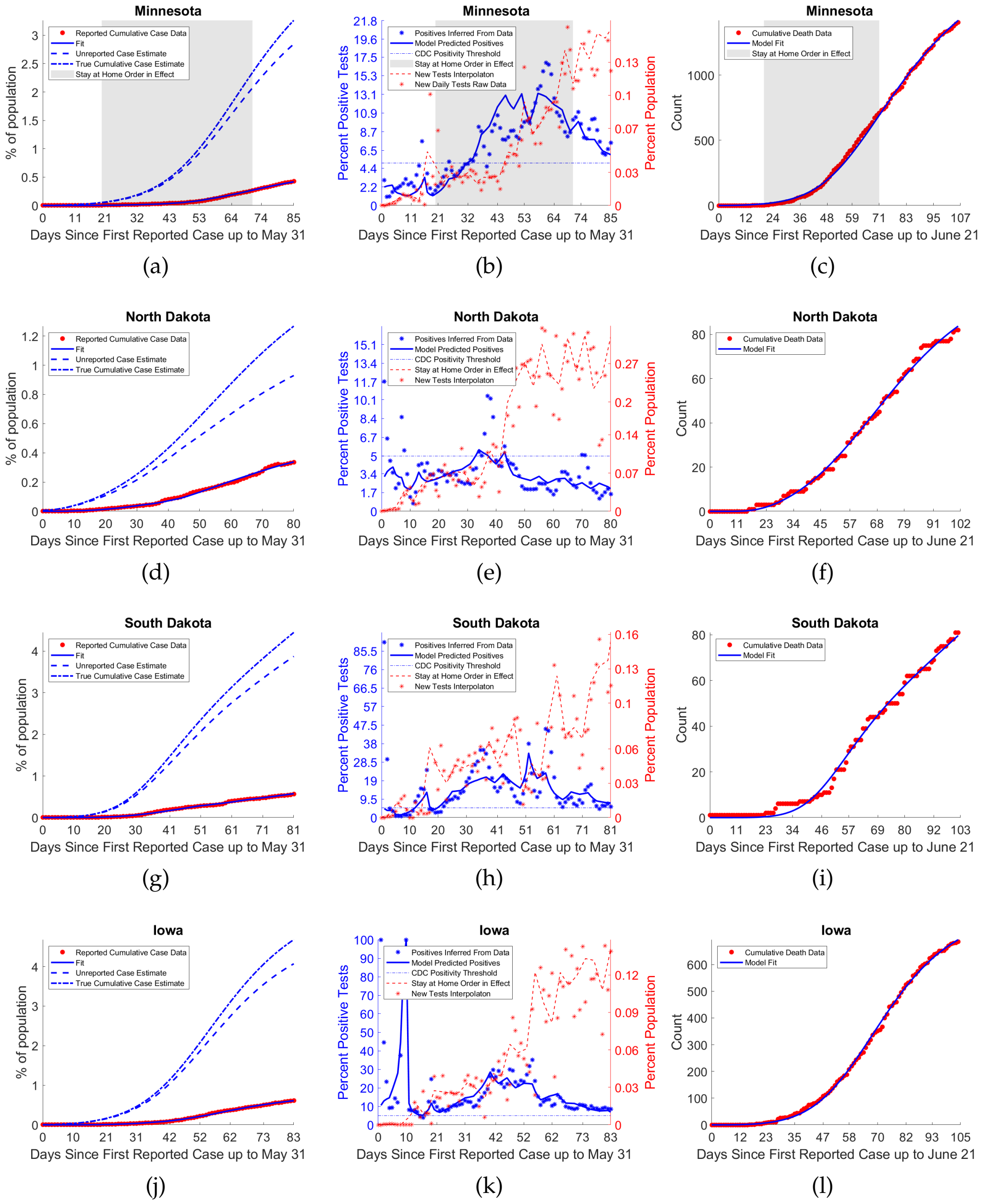


Figure S7: Fits for states and territories in the Midwest. States and territories in this region include Michigan, Wisconsin, Ohio, Indiana, Illinois, Minnesota, North Dakota, South Dakota, Iowa, Nebraska, Kansas, and Missouri. Figure 2 of 3. See also figures S11,S8

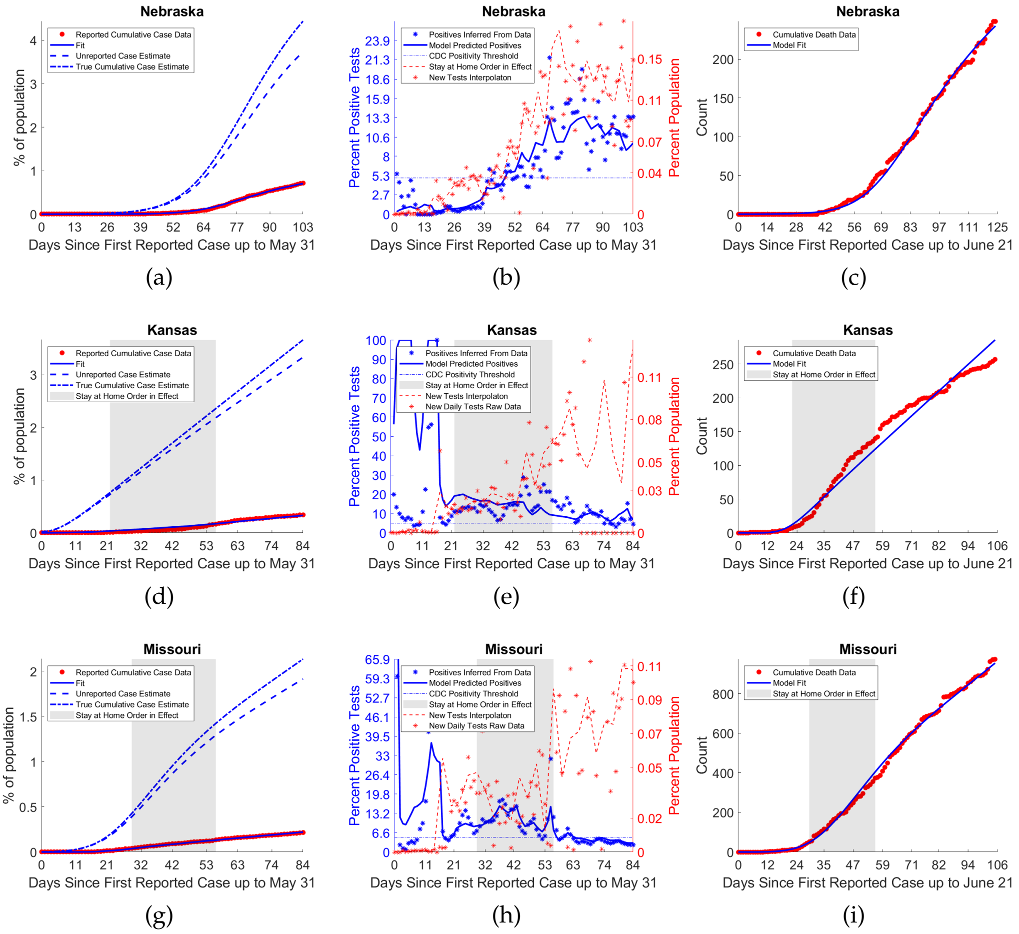


Figure S8: Fits for states and territories in the Midwest. States and territories in this region include Michigan, Wisconsin, Ohio, Indiana, Illinois, Minnesota, North Dakota, South Dakota, Iowa, Nebraska, Kansas, and Missouri. Figure 3 of 3See also figures S11,S12

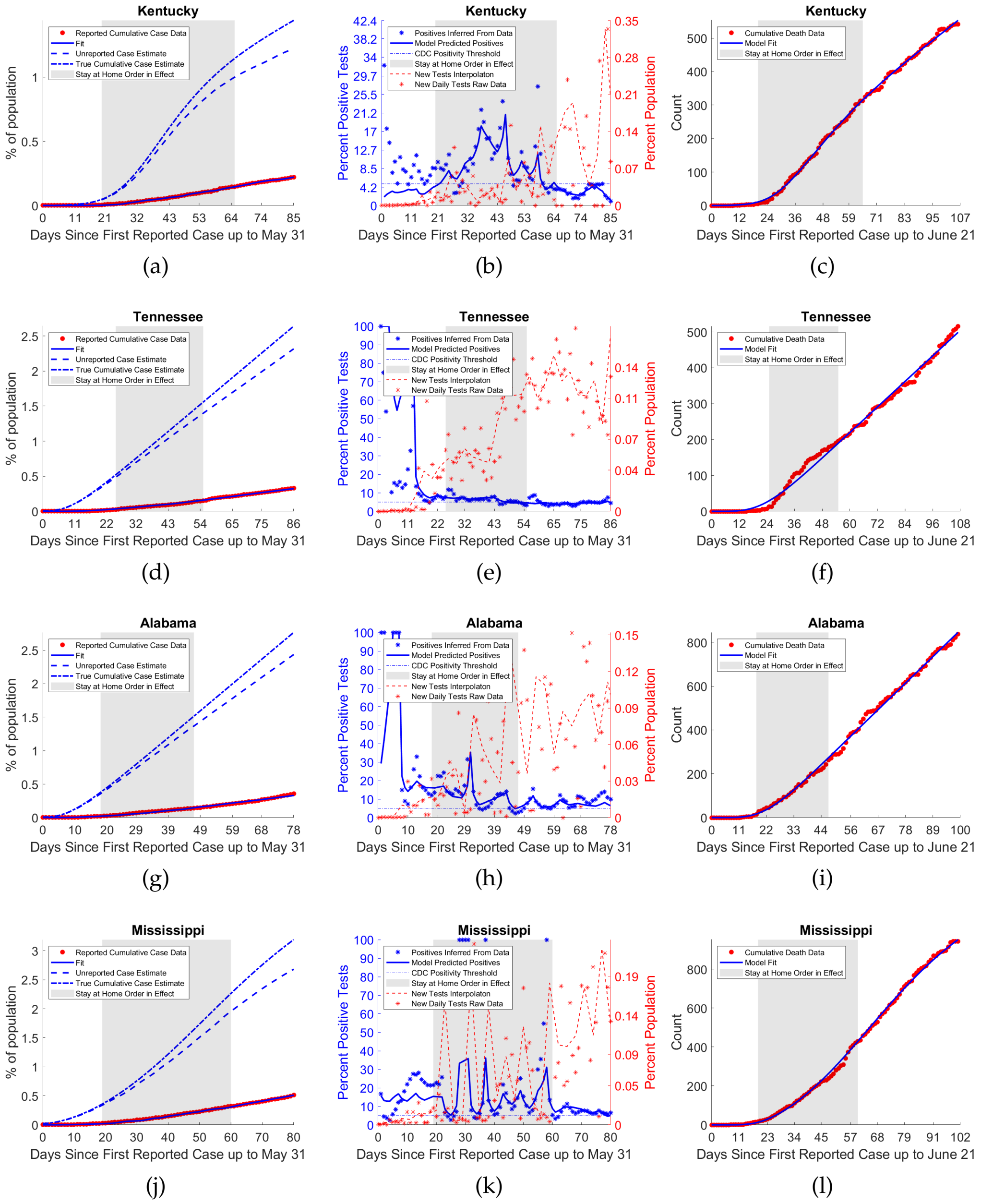


Figure S9: Fits for states and territories in the Central South. States and territories in this region include Kentucky, Tennessee, Alabama, Mississippi, Oklahoma, Arkansas, Texas, and Louisiana. Figure 1 of 2. See also figure S10

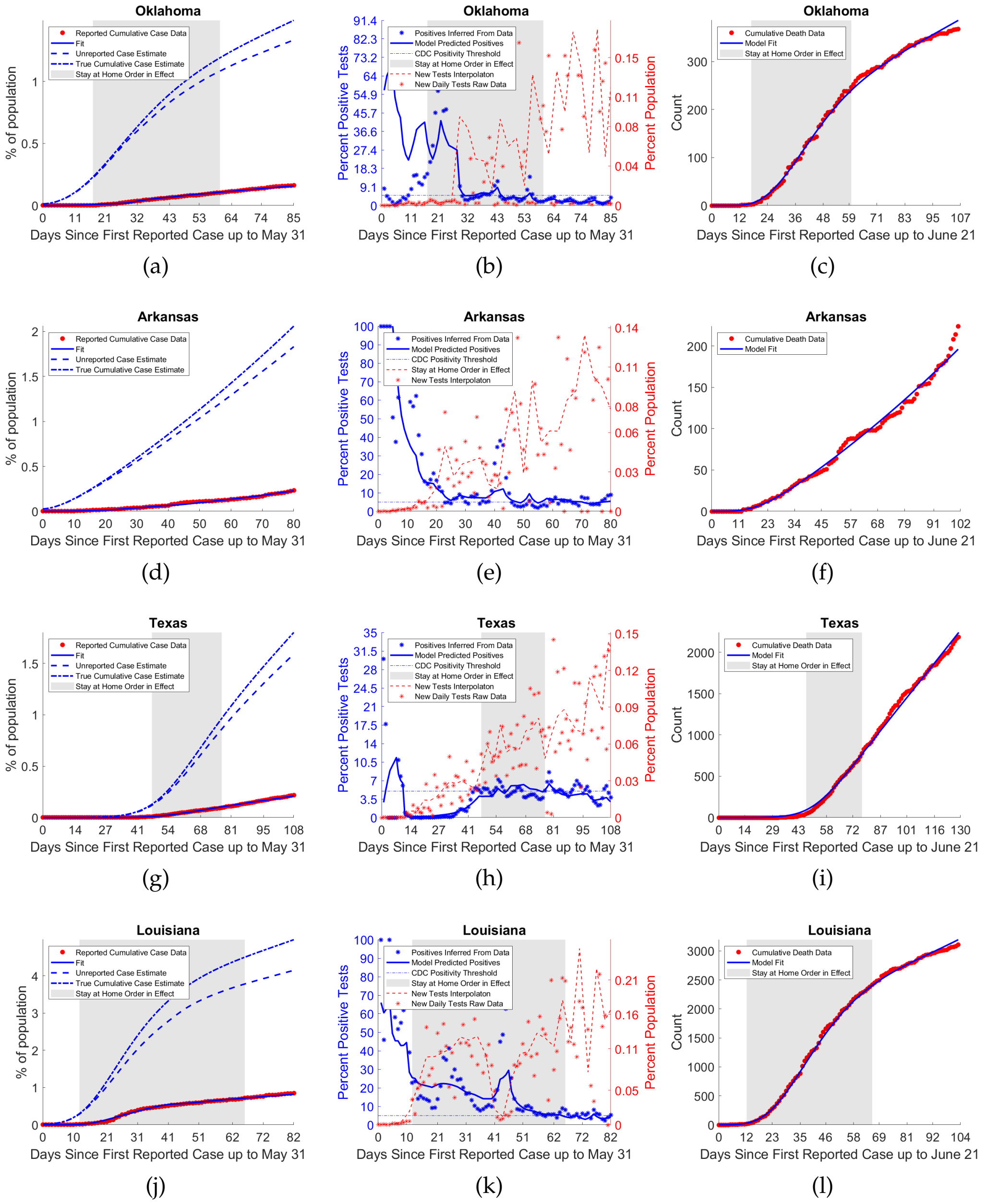


Figure S10: Fits for states and territories in the Central South. States and territories in this region include Kentucky, Tennessee, Alabama, Mississippi, Oklahoma, Arkansas, Texas, and Louisiana. Figure 2 of 2 See also figure S9

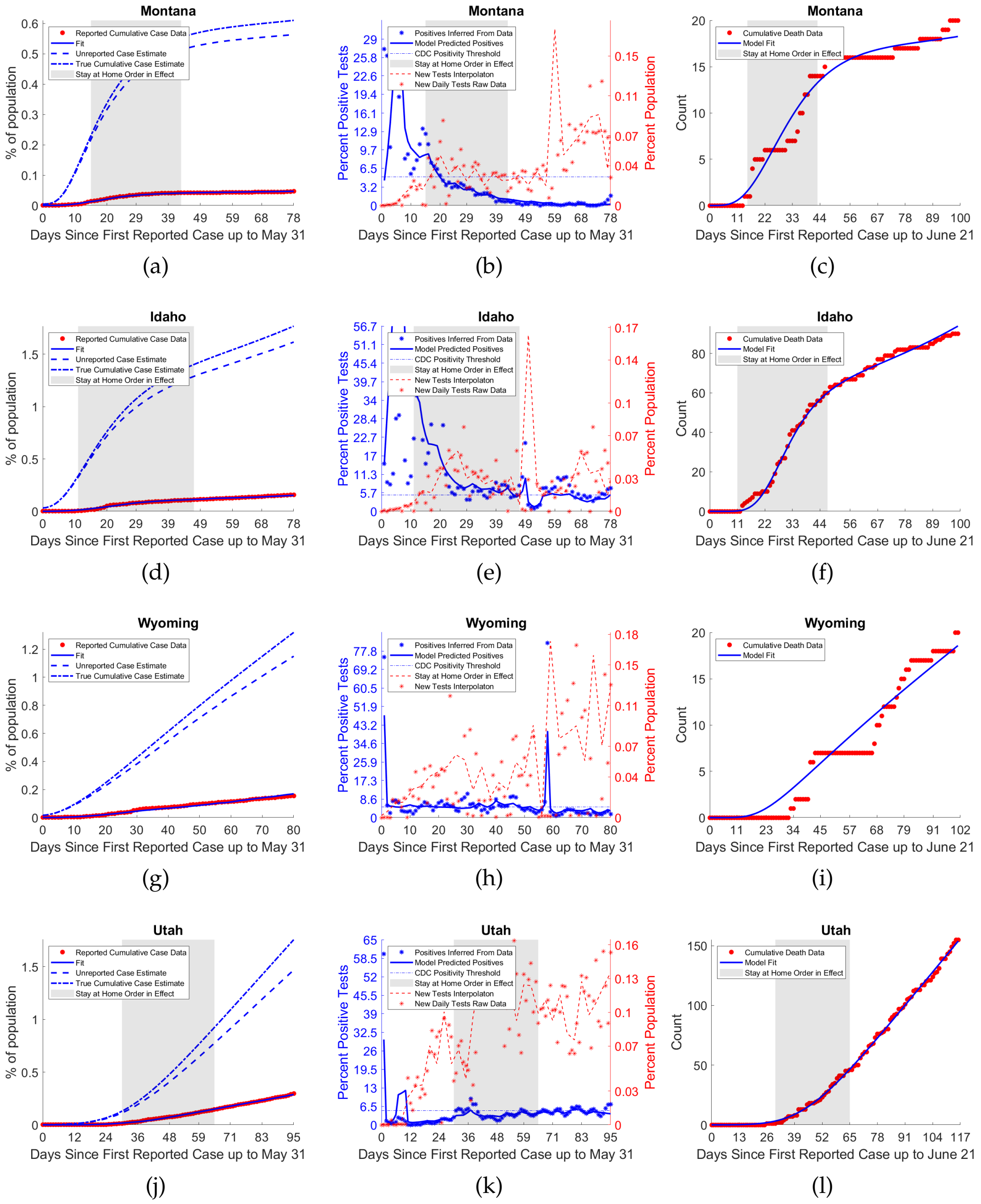

Figure S11: Fits for states and territories in the Mountain West. States and territories in this region include Montana, Idaho, Wyoming, Utah, Nevada, Arizona, New Mexico, and Colorado. Figure 1 of 2 See also figure S12

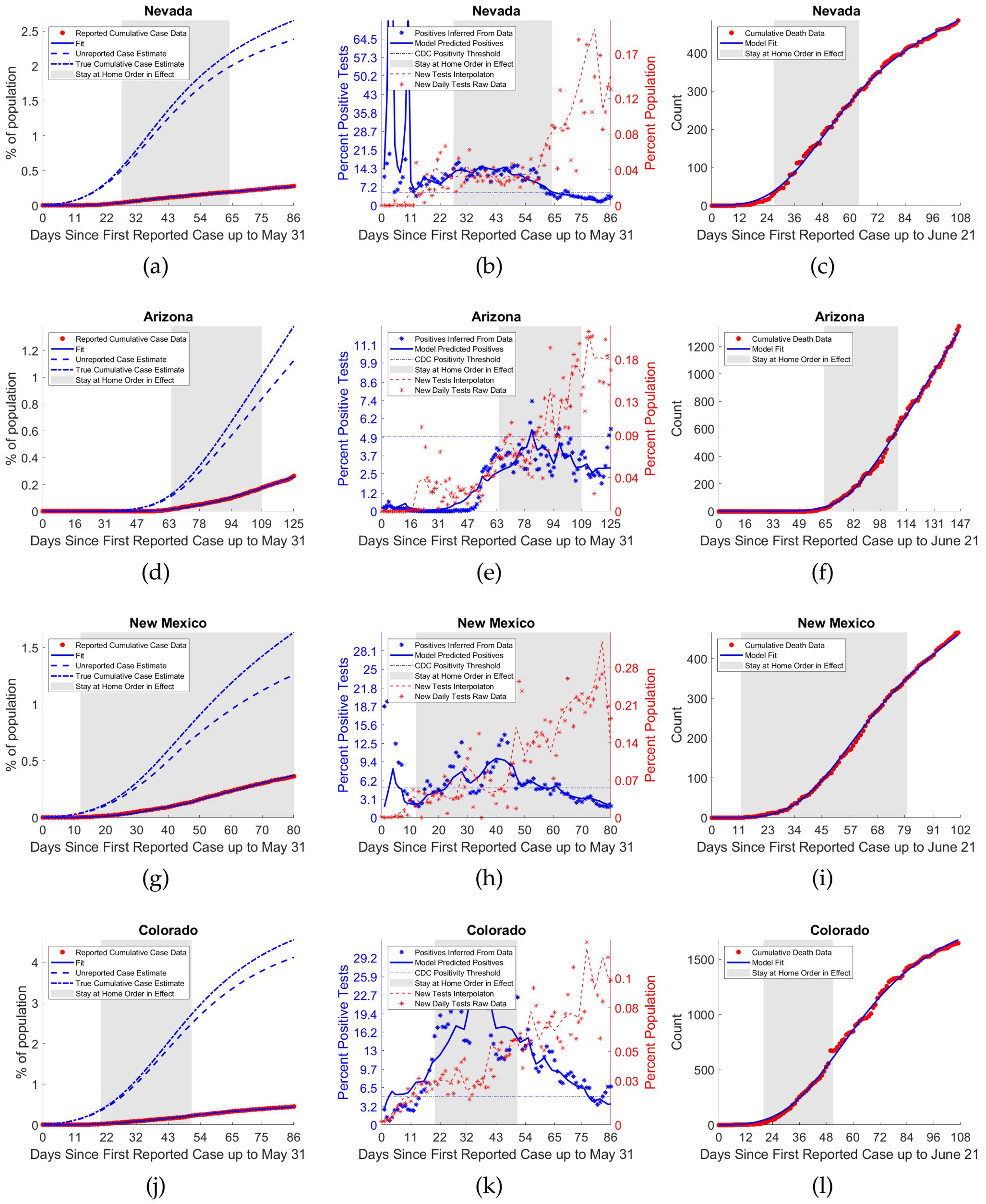


Figure S12: Fits for states and territories in the Mountain West. States and territories in this region include Montana, Idaho, Wyoming, Utah, Nevada, Arizona, New Mexico, and Colorado. Figure 2 of 2 See also figure S11

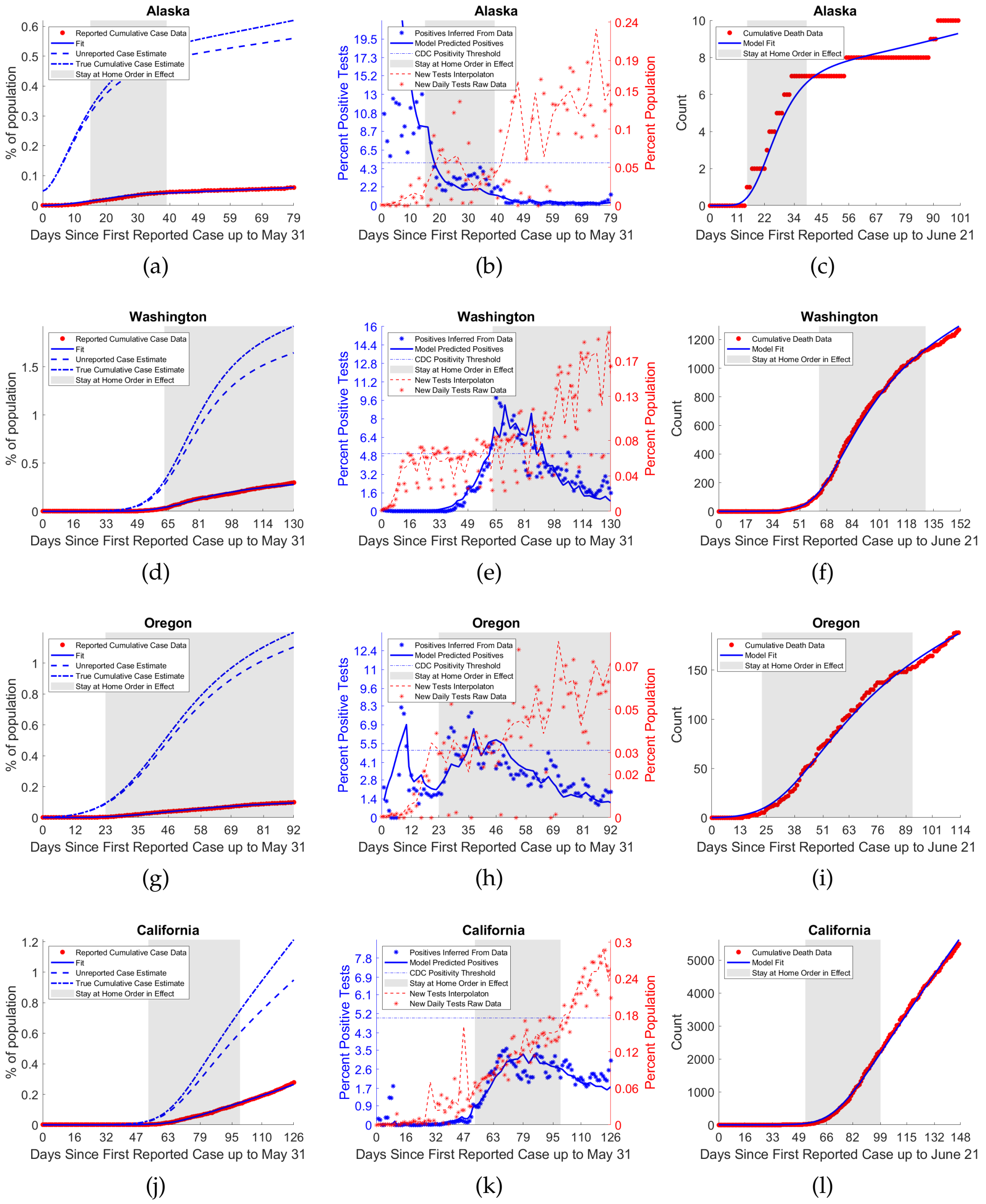


Figure S13: Fits for states and territories in the Pacific West. States and territories in this region include Alaska, Washington, Oregon, California, Hawaii, Guam, and the Northern Marianas Islands. Figure 1 of 2 See also figure S14

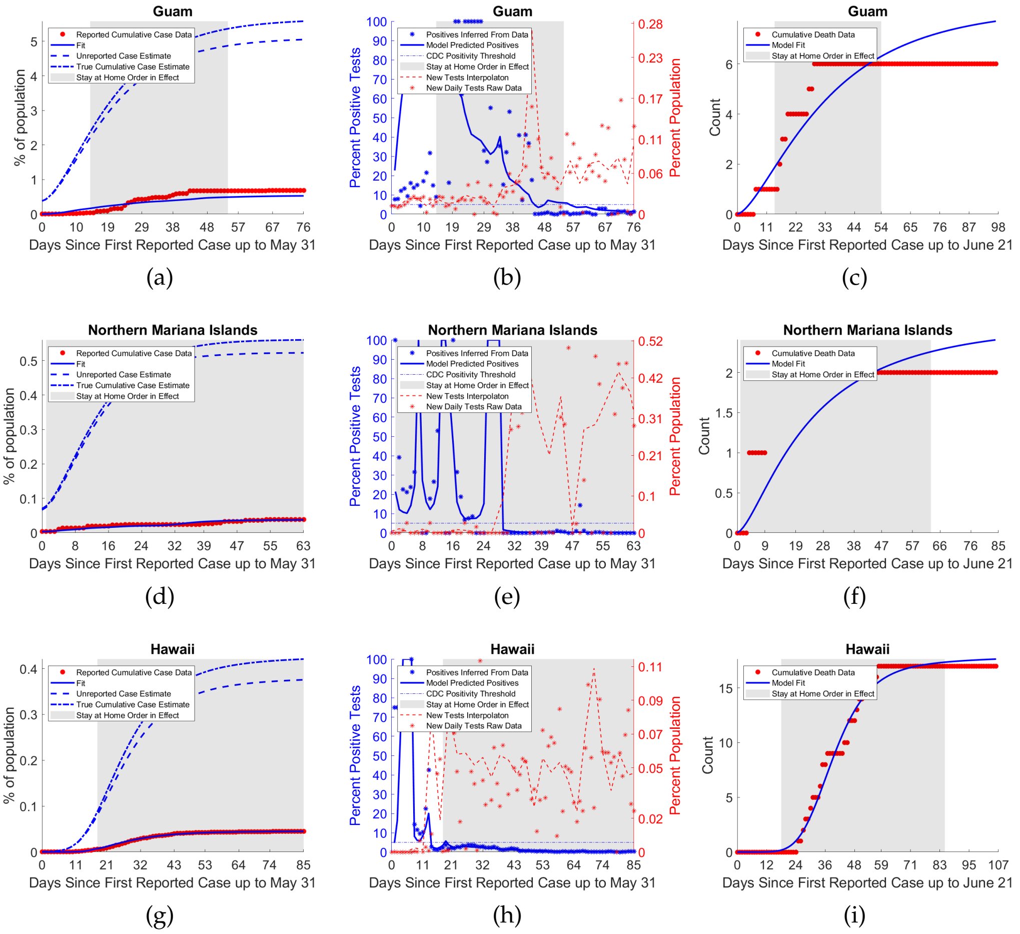


Figure S14: Fits for states and territories in the Pacific West. States and territories in this region include Alaska, Washington, Oregon, California, Hawaii, Guam, and the Northern Marianas Islands. Figure 2 of 2. See also figure S13

### 2. Model Robustness

In order to test the robustness of parameter results, particularly those of IFR, ratio of True to Reported Cases, positivity, and rho, we considered two alternative scenarios. First, we fixed the IFR, model parameter, $\xi = 0.0086$, at the fit value for the United States as a whole, for all states and territories, while fitting the other model parameters as reported in the main text. Second we fixed $A/N$ and $k$ at the fit value for the United States and carried out the same process again (see table S2 for specific values).

As can be seen in both figures S15,S16 and table S1 both of these alternative fitting procedures produced significantly increased residual values, uniformly across states in the first case, with increasing effect as the fixed parameters in question differed more significantly from those provided in the main body of the text, with qualitatively most fittings experiencing significant decrease in visual quality of fit to the underlying data in the first alternative fitting, and badly so in a number of states in the second.

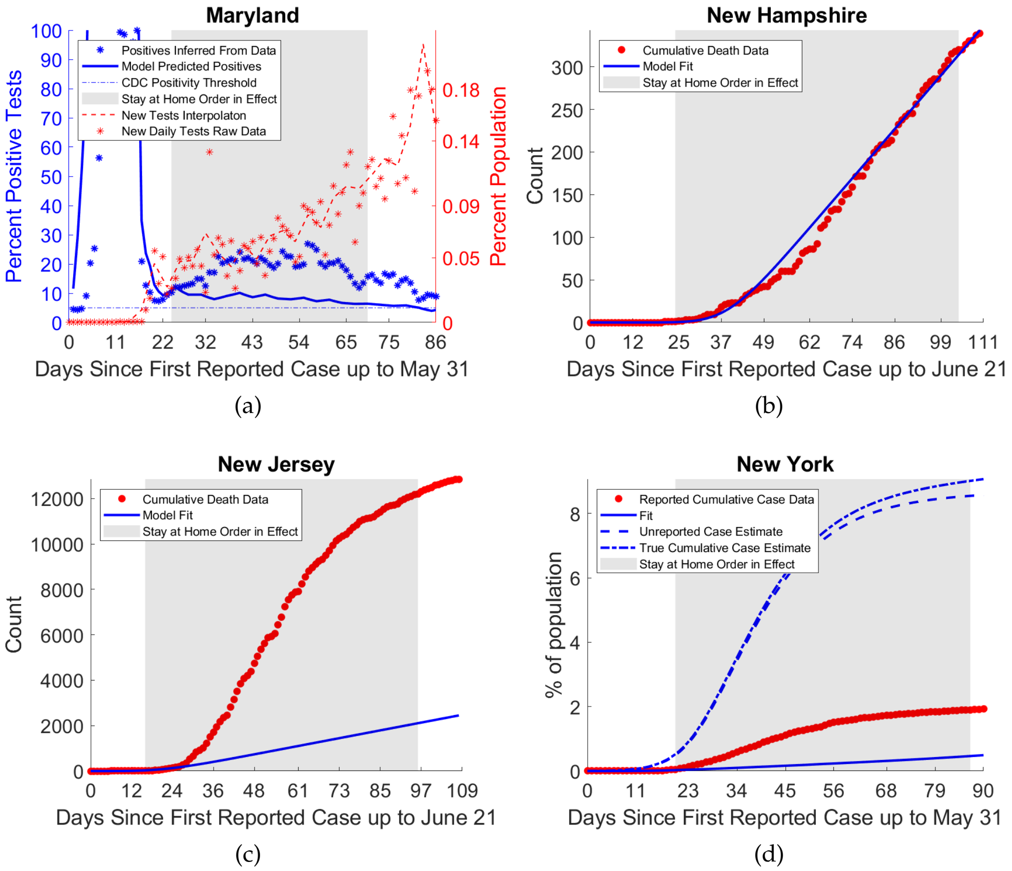


Figure S15: (a), (c), (d) are examples of the poor quality of fit for most states under alternative fitting procedure 1, with IFR fixed at 0.086. (b) shows that even for New Hampshire, which had the 3rd lowest increase in residual relative to baseline in this scenario, still has visibly worse fit for cumulative deaths compared to baseline.

Thus we conclude that our model is not over-identified, and that we need the flexibility of fitting both $\rho$ and $\xi$ in order to obtain good fits across all states and territories.

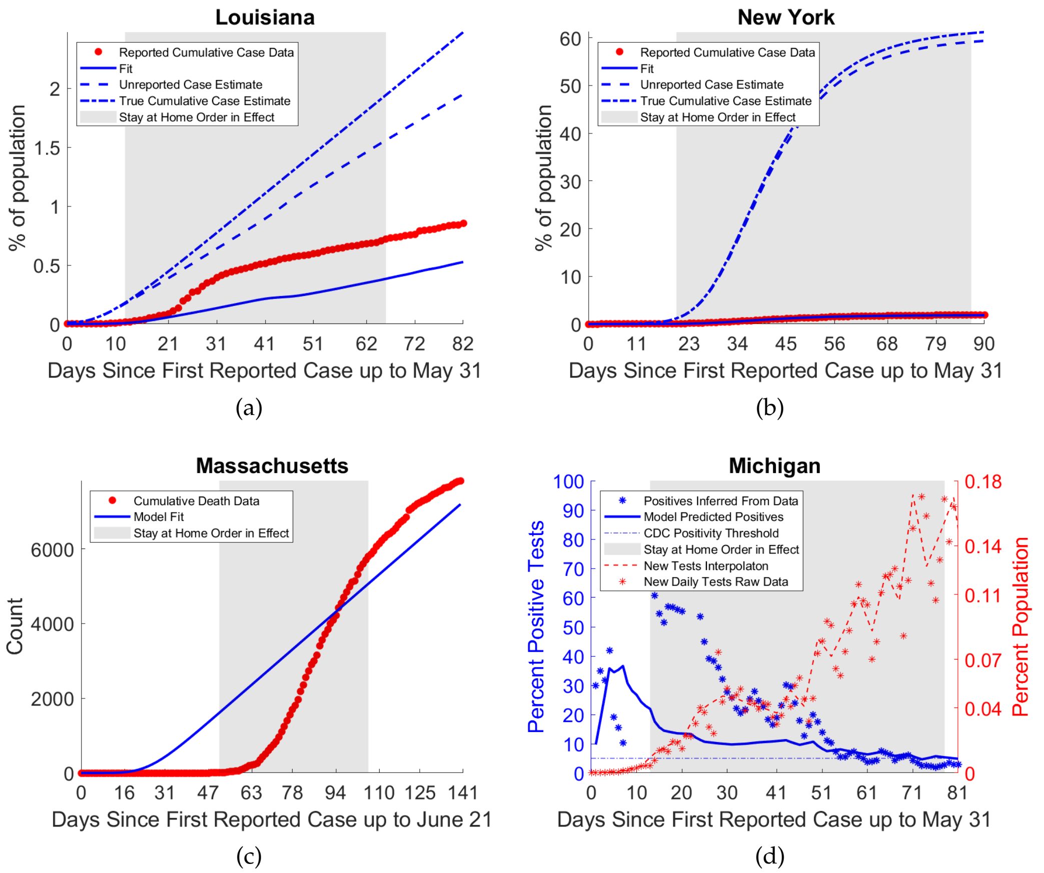


Figure S16: (a)-(d) are examples of the poor quality of fit for many states under alternative fitting procedure 2, with $\rho$ fixed at its fit parameter values for the aggregated US fit. Though some states saw moderate decreases in residual other states saw large magnitude increases in error.

Table S1: Comparison of residual across baseline fitting and two considered alternative fitting scenarios. The results suggest that our model is not over identified.

| State | Baseline Residual | Alt. Fitting 1 Change in Residual (%) | Alt Fitting 2 Change in Residual (%) |
|---|---|---|---|
| Alabama | 4.206345e+04 | 915.631541 | 10.890491 |
| Alaska | 4.615980e+01 | 4471.896847 | -3.042317 |
| Arizona | 4.657744e+04 | 8.630493 | 0.236204 |
| Arkansas | 6.367533e+03 | 3994.041526 | 31.393464 |
| California | 1.048639e+06 | 108.679742 | 33.684916 |
| Colorado | 9.923768e+04 | 5664.232285 | -15.861060 |
| Connecticut | 6.631159e+05 | 99722.714230 | 21950.764709 |
| Delaware | 3.663345e+03 | 18856.640189 | 10.249598 |
| District of Columbia | 1.728646e+04 | 1371.605940 | 4.647304 |
| Florida | 2.756271e+05 | 7824.432291 | 18.560736 |
| Georgia | 1.967957e+05 | 8024.669882 | 15.070394 |
| Guam | 2.259009e+02 | 496.804326 | -22.807001 |
| Hawaii | 6.527919e+01 | 12262.851571 | -2.857237 |
| Idaho | 8.588132e+02 | 6704.486143 | -10.383475 |
| Illinois | 1.193061e+06 | 23382.169701 | 7.484186 |
| Indiana | 2.557395e+05 | 19233.185115 | -11.801250 |
| Iowa | 1.093170e+04 | 9550.595539 | 28.805513 |
| Kansas | 5.221363e+04 | 1319.187171 | -22.398653 |
| Kentucky | 5.599718e+03 | 0.409305 | 0.014628 |
| Louisiana | 4.869764e+05 | 53355.284441 | 17338.863741 |
| Maine | 2.487835e+03 | 453.737408 | 13.076172 |
| Maryland | 4.326181e+05 | 22879.178371 | 8.549645 |
| Massachusetts | 6.717183e+05 | 239802.995245 | 96611.534355 |
| Michigan | 6.767791e+05 | 173954.418266 | 57630.223640 |
| Minnesota | 6.164779e+04 | 6317.489951 | -10.505335 |
| Mississippi | 2.465630e+04 | 1307.620183 | 49.563402 |
| Missouri | 4.072829e+04 | 4317.638394 | 49.661619 |
| Montana | 1.285880e+02 | 5620.125651 | 2.484751 |
| Nebraska | 3.212663e+03 | 9692.020053 | 1.746522 |
| Nevada | 5.054666e+03 | 11406.721872 | 0.347801 |
| New Hampshire | 2.182800e+04 | 6.241721 | 10.294945 |
| New Jersey | 2.698465e+06 | 288705.623915 | 56204.747302 |
| New Mexico | 3.810991e+03 | 55.671747 | -53.193424 |
| New York | 1.582585e+08 | 38495.440996 | 0.697325 |
| North Carolina | 3.253540e+04 | 3673.236007 | -18.143785 |
| North Dakota | 4.461594e+02 | 121.118898 | -6.918899 |
| Northern Mariana Islands | 2.443229e+01 | 5195.516351 | -2.669997 |
| Ohio | 1.483507e+05 | 13721.418418 | 62.480528 |
| Oklahoma | 9.237180e+03 | 169.578393 | -14.575337 |
| Oregon | 3.400838e+03 | 4495.139675 | 8.374007 |
| Pennsylvania | 6.291545e+05 | 143132.682898 | 38694.621036 |
| Puerto Rico | 9.349543e+03 | 992.434233 | 29.707681 |
| Rhode Island | 2.729352e+04 | 4506.704650 | -18.076879 |
| South Carolina | 3.471895e+04 | 4437.531351 | 30.312198 |
| South Dakota | 6.862404e+02 | 10908.291363 | 7.447183 |
| Tennessee | 2.093300e+04 | 12212.549844 | -21.656223 |
| Texas | 2.849479e+05 | 3513.104570 | -6.634123 |
| Utah | 6.628265e+02 | 29480.415490 | 3.695024 |
| Vermont | 1.546140e+03 | 98.546912 | -6.880094 |
| Virgin Islands | 3.363363e+01 | 331.924473 | 9.230913 |
| Virginia | 1.408870e+05 | 9318.656840 | 3.265786 |
| Washington | 1.583526e+05 | 10.157508 | 7.717952 |
| West Virginia | 1.250249e+03 | 2523.426439 | 18.722583 |
| Wisconsin | 7.915386e+04 | 6.023655 | 15.007094 |
| Wyoming | 2.957362e+02 | 1576.298324 | -2.311043 |

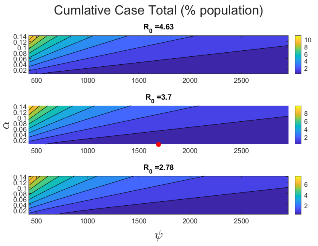


(a)

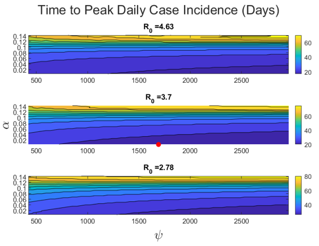


(b)

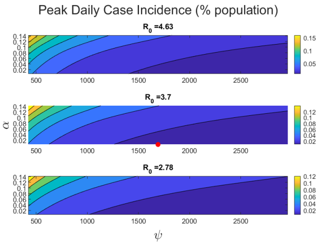


(c)

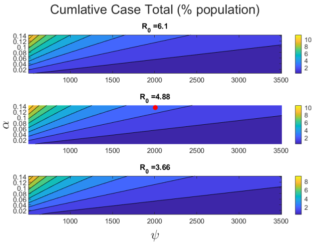


(d)

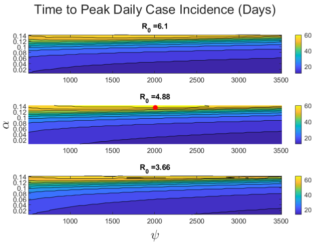


(e)

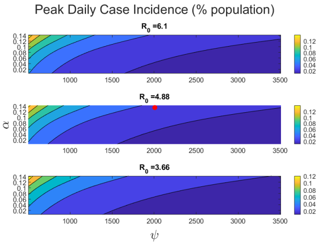


(f)

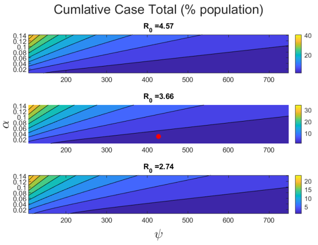


(g)

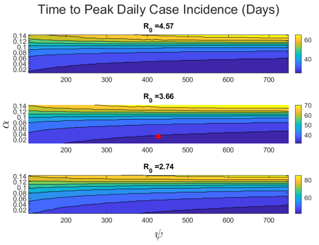


(h)

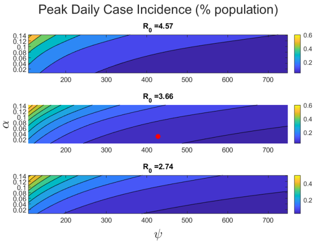


(i)

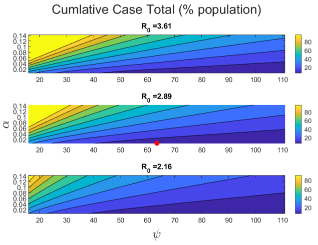


(j)

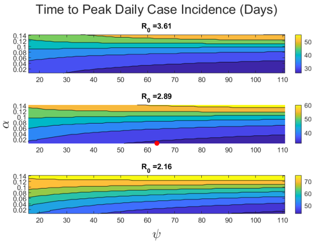


(k)

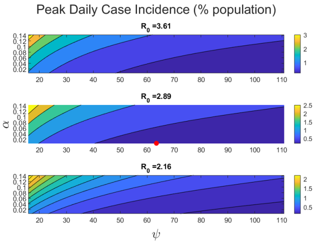


(l)

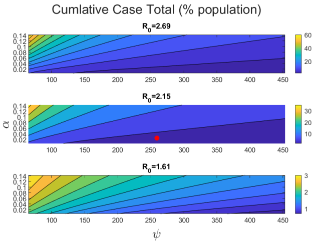


(m)

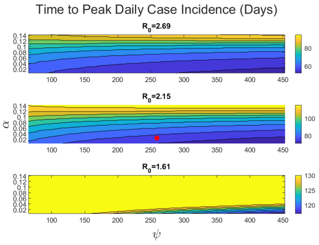


(n)

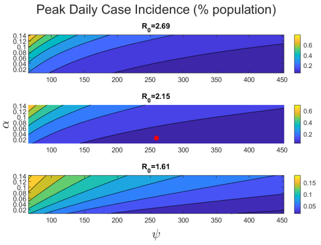


(o)

Figure S17: Examination of the combined effects of $\mathcal{R}_0$,$\psi$, $\alpha$ on different aspects of epidemic trajectory Row 1: Hawaii, Row 2:Tennessee, Row 3: Georgia, Row4: New York, Row5: The United States

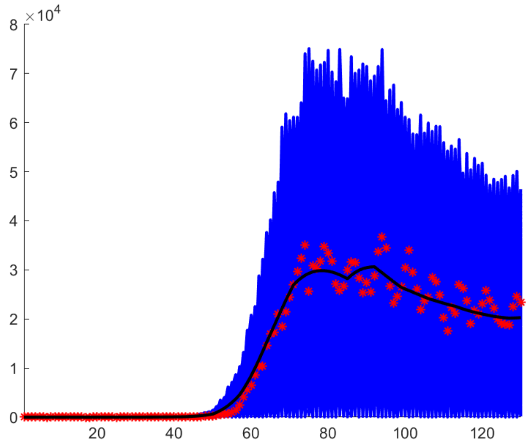


Figure S18: Synthetic covering data-sets (in blue), together with original data (red), and baseline daily case curve inferred from cumulative case fit (black) that were used for United states fit confidence interval generation (60% noise level).

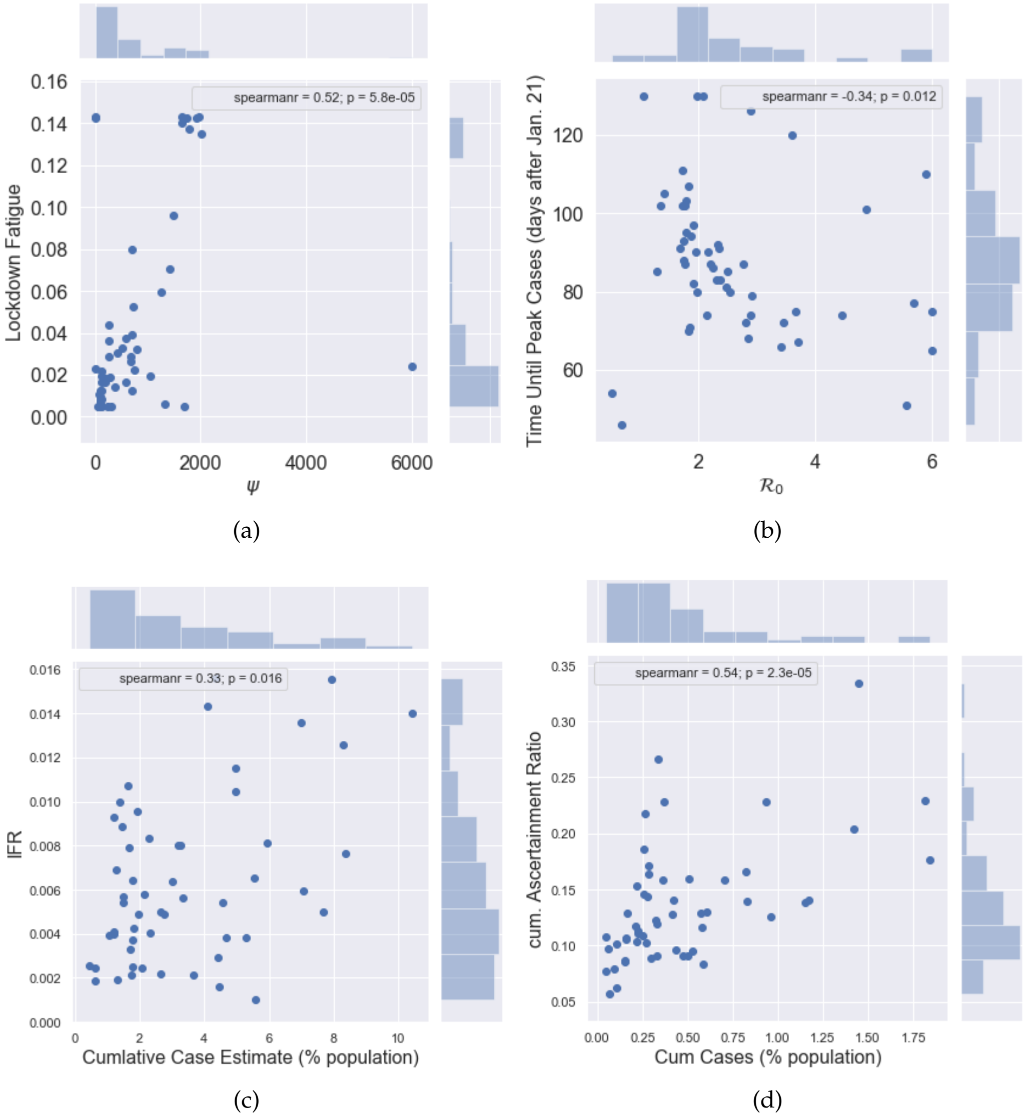


Figure S19: Correlation plots not included in the main body of the text.

Table S2: Parameters and quantities not included in main body of the paper. Deaths are as of June 21st.

| State | $k$ | $A$ | $\eta$ | $\mu/\eta$ | Deaths (%) |
|---|---|---|---|---|---|
| Alabama | 0.203 | 2967.1 | 6.07 | 3.46 | 0.017 |
| Alaska | 0.323 | 675.5 | 12.00 | 1.75 | 0.001 |
| Arizona | 0.293 | 6630.7 | 12.00 | 1.75 | 0.018 |
| Arkansas | 0.188 | 1622.9 | 5.34 | 3.93 | 0.006 |
| California | 0.406 | 68667.8 | 1.71 | 12.26 | 0.014 |
| Colorado | 0.169 | 4151.9 | 2.47 | 8.49 | 0.029 |
| Connecticut | 0.285 | 4361.1 | 3.18 | 6.61 | 0.122 |
| Delaware | 0.240 | 1001.0 | 2.47 | 8.49 | 0.045 |
| District of Columbia | 0.260 | 785.9 | 12.00 | 1.75 | 0.076 |
| Florida | 0.260 | 15940.8 | 3.05 | 6.88 | 0.014 |
| Georgia | 0.287 | 8705.9 | 1.71 | 12.30 | 0.024 |
| Guam | 0.378 | 178.8 | 0.50 | 42.00 | 0.005 |
| Hawaii | 0.153 | 617.3 | 9.52 | 2.21 | 0.001 |
| Idaho | 0.228 | 1165.6 | 11.92 | 1.76 | 0.005 |
| Illinois | 0.375 | 20383.8 | 0.50 | 42.00 | 0.054 |
| Indiana | 0.134 | 3851.8 | 0.81 | 25.97 | 0.038 |
| Iowa | 0.186 | 1680.3 | 4.09 | 5.13 | 0.022 |
| Kansas | 0.168 | 1607.5 | 12.00 | 1.75 | 0.010 |
| Kentucky | 0.467 | 5963.4 | 0.50 | 42.00 | 0.012 |
| Louisiana | 0.358 | 6596.6 | 2.89 | 7.26 | 0.069 |
| Maine | 0.468 | 2698.0 | 12.00 | 1.75 | 0.008 |
| Maryland | 0.300 | 7783.6 | 0.82 | 25.56 | 0.052 |
| Massachusetts | 0.357 | 7152.9 | 11.68 | 1.80 | 0.115 |
| Michigan | 0.240 | 6852.8 | 4.22 | 4.98 | 0.063 |
| Minnesota | 0.219 | 5282.8 | 0.50 | 42.00 | 0.025 |
| Mississippi | 0.316 | 2685.9 | 12.00 | 1.75 | 0.032 |
| Missouri | 0.152 | 2666.7 | 4.33 | 4.85 | 0.016 |
| Montana | 0.248 | 1134.6 | 2.95 | 7.12 | 0.002 |
| Nebraska | 0.248 | 2057.2 | 0.50 | 42.00 | 0.013 |
| Nevada | 0.277 | 3655.6 | 2.26 | 9.30 | 0.016 |
| New Hampshire | 0.259 | 1008.2 | 11.49 | 1.83 | 0.025 |
| New Jersey | 0.672 | 17047.2 | 3.18 | 6.60 | 0.143 |
| New Mexico | 0.461 | 2761.9 | 12.00 | 1.75 | 0.022 |
| New York | 0.430 | 41826.2 | 1.60 | 13.09 | 0.164 |
| North Carolina | 0.231 | 10395.8 | 12.00 | 1.75 | 0.012 |
| North Dakota | 0.451 | 982.9 | 12.00 | 1.75 | 0.011 |
| N. Marianas | 0.697 | 169.8 | 0.50 | 42.00 | 0.004 |
| Ohio | 0.114 | 3793.5 | 3.35 | 6.26 | 0.023 |
| Oklahoma | 0.250 | 3338.6 | 12.00 | 1.75 | 0.010 |
| Oregon | 0.114 | 2061.2 | 0.50 | 42.00 | 0.004 |
| Pennsylvania | 0.107 | 5843.0 | 7.72 | 2.72 | 0.051 |
| Puerto Rico | 0.595 | 8144.4 | 0.50 | 42.00 | 0.005 |
| Rhode Island | 0.503 | 1523.7 | 2.69 | 7.81 | 0.085 |
| South Carolina | 0.216 | 3179.6 | 3.90 | 5.38 | 0.012 |
| South Dakota | 0.192 | 485.9 | 11.58 | 1.81 | 0.009 |
| Tennessee | 0.212 | 6215.2 | 4.63 | 4.53 | 0.007 |
| Texas | 0.201 | 25019.0 | 0.50 | 42.00 | 0.008 |
| Utah | 0.219 | 2010.0 | 3.41 | 6.15 | 0.005 |
| Vermont | 0.257 | 565.5 | 5.91 | 3.55 | 0.009 |
| Virgin Islands | 0.091 | 41.6 | 12.00 | 1.75 | 0.006 |
| Virginia | 0.163 | 5973.8 | 0.56 | 37.46 | 0.019 |
| Washington | 0.279 | 8961.4 | 0.50 | 41.99 | 0.017 |
| West Virginia | 0.263 | 2017.8 | 7.22 | 2.91 | 0.005 |
| Wisconsin | 0.314 | 7845.8 | 0.50 | 42.00 | 0.013 |
| Wyoming | 0.242 | 400.9 | 9.75 | 2.15 | 0.003 |
| United States | 0.358 | 2.71e+5 | 12.00 | 1.75 | 0.099 |
